# Integrating explainable machine learning and transcriptomics data reveals cell-type specific immune signatures underlying macular degeneration

**DOI:** 10.1101/2024.10.26.24316189

**Authors:** Khang Ma, Hosei Nakajima, Nipa Basak, Arko Barman, Rinki Ratnapriya

**Affiliations:** Department of Ophthalmology, Baylor College of Medicine, 6565 Fannin St, NC205, Houston, TX, 77030 United States; Data to Knowledge Lab, Rice University, Houston, TX, 77005, United States; Department of Electrical & Computer Engineering, Rice University, Houston, TX, 77005, United States; Department of Statistics, Rice University, Houston, TX, 77005, United States; Department of Biochemistry and Molecular Pharmacology, Baylor College of Medicine, 6565 Fannin St, NC205, Houston, TX, 77030 United States

## Abstract

Genome-wide association studies (GWAS) have established a key role of dysfunctional immune response in the etiology of Age-related Macular Degeneration (AMD). However, immune cells constitute a small proportion of the retina, and their role in AMD is not completely resolved. Here we develop an explainable machine learning pipeline using transcriptome data 453 donor retinas, identifying 81 genes distinguishing AMD from controls with an AUC-ROC of 0.80 (CI 0.70-0.92). These genes show enrichment for pathways involved in immune response, complement and extracellular matrix and connected to known AMD genes through co-expression networks and gene expression correlation. The majority of these genes were enriched in their expression within retinal glial cells, particularly microglia and astrocytes. Their role in AMD was further strengthened by cellular deconvolution, which identified distinct differences in microglia and astrocytes between normal and AMD. We corroborated these findings using independent single-cell data, where several of these candidate genes exhibited differential expression. Finally, the integration of AMD-GWAS data identified a common regulatory variant, rs4133124 at *PLCG2*, as a novel AMD-association. Collectively, our study provides molecular insights into the recurring theme of immune dysfunction in AMD and highlights the significance of glial cell differences as an important determinant of AMD progression.

## INTRODUCTION

Variation in gene expression has emerged as a significant source of phenotypic diversity among individuals and populations^1^. Additionally, human genetic studies have highlighted the critical role of gene expression dysregulation in both rare^2^ and common^3^ diseases. Understanding the dysregulation of gene expression in different diseases is essential for deciphering the underlying molecular mechanisms and identifying potential targets for therapeutic intervention. The cellular context has a profound influence on gene expression and regulation, emphasizing the importance of comprehensively studying transcriptome regulation in disease-relevant cells and tissues. However, the availability of disease-relevant tissues in a large number of individuals presents a significant challenge. Additionally, gene expression in humans is influenced by genetic variants, epigenetic changes, environmental factors, or a combination of these factors^4^ making gene expression studies uniquely challenging to identify consistent disease-related patterns.

Age-related Macular Degeneration (AMD) is the leading cause of irreversible vision loss in people over 50 years of age^5^. It is a neurodegenerative disease that afflicts almost 10 million individuals in the United States alone and this number is expected to double by 2050^6^. AMD results from the deterioration of the photoreceptor support system, which includes the retinal pigment epithelium (RPE), Bruch’s membrane (BrM), and the choroidal vasculature, leading to the death of photoreceptors primarily in the central region of the retina called macula^7^. It is a complex, multifactorial disease that is caused by the cumulative impact of genetic predisposition, environmental stress and late aging^8^. Knowledge of genetic risk factors underlying AMD susceptibility has advanced rapidly with the advent of Genome-wide Association Studies (GWAS), which have successfully identified 52 independent genetic variants at 34 loci^9^ establishing a strong genetic component of AMD that is mostly driven by common variants^10, 11^. These findings have implicated immune, complement, cholesterol and lipid metabolism, extracellular/collagen matrix, and angiogenesis pathways in AMD pathogenesis^9^. Among them, variants identified in complement and immunoinflammatory genes such as *CFH, CFI* and *C3* have become the essential core for AMD genetics because of high effect size associated with these genes^9, 10, 11^. Additionally, substantial clinical evidence underscores the significant involvement of immunologic processes such as the production of inflammatory molecules, recruitment of macrophages, complement activation and microglial activation in AMD pathology^12^. The majority of AMD-associated variants reside in the non-coding region of the genome mediate the disease risk through gene expression regulation in retina^13, 14^ and RPE^15^. However, the molecular mechanisms underlying AMD, especially the cellular vulnerability, are poorly understood.

Recent advancements in genomics have transformed biomedical research into digitalized, data-intensive science that has broadened its application in biology and medicine. However, the scale, complexity and high information content are significant barriers in its application. These limitations have encouraged the application of machine learning (ML) methods to help make informed decisions to drive novel biological hypotheses and translate them into tangible therapeutics^16, 17^. In particular, ML-based approaches have been frequently used to obtain insights related to regulatory regions of the genome and how they impact gene expression and phenotypic changes^18^. Within ophthalmology, ML has occupied a niche based on studies of retinal fundus and optical coherence tomography (OCT) images and visual fields by achieving robust diagnosis performance in detecting various disease including diabetic retinopathy, retinopathy of prematurity, glaucoma, macular edema and AMD^19^.

Comparative transcriptome studies in disease-relevant tissues and cell-types hold great potential for identifying new genes as well as investigating mechanisms underlying the disease. However, small sample sizes and the high heterogeneity of study samples impose significant challenges in its interpretation. ML-based feature selection offers a great tool to address these limitations. Here we present the development of explainable ML models to classify the AMD based on their expression profiles of 453 samples. To the best of our knowledge, this is the first study to rigorously test gene expression data for their ability to accurately distinguish AMD from normal. We further analyzed the features selected for ML models using pathways and co-expression regulation networks. Finally, we integrate the data from AMD-GWAS and single-cell transcriptomics to identify the genes and cell types associated with AMD pathology.

## RESULTS

### Feature selection and machine learning model reveals a core set of 81 AMD genes

Transcriptome data often suffers from the “curse of dimensionality” as tens of thousands of genes can be profiled in a single RNAseq experiment vs the limited number of subjects. Thus, we developed a pipeline (**Fig. 1A**) to reduce the dimension and improve the efficiency and interpretability of downstream analyses. We implemented three feature selection methods, ANOVA (analysis of variance) F-test, AUC (area under the curve), and Kruskal-Wallis test to identify the most relevant features. We divided the dataset into an 80% training set and a 20% testing set. We used the training set to identify the most influential features within the training data and evaluated the model’s performance on the separate 20% testing data, employing appropriate evaluation metrics. Comparing the features of top the 100 features identified across 1000 iterations selected by each method, we identified 81 genes (referred as ML-genes) that were common across three methods (**Supplementary Fig. 1**).

**Figure 1:**
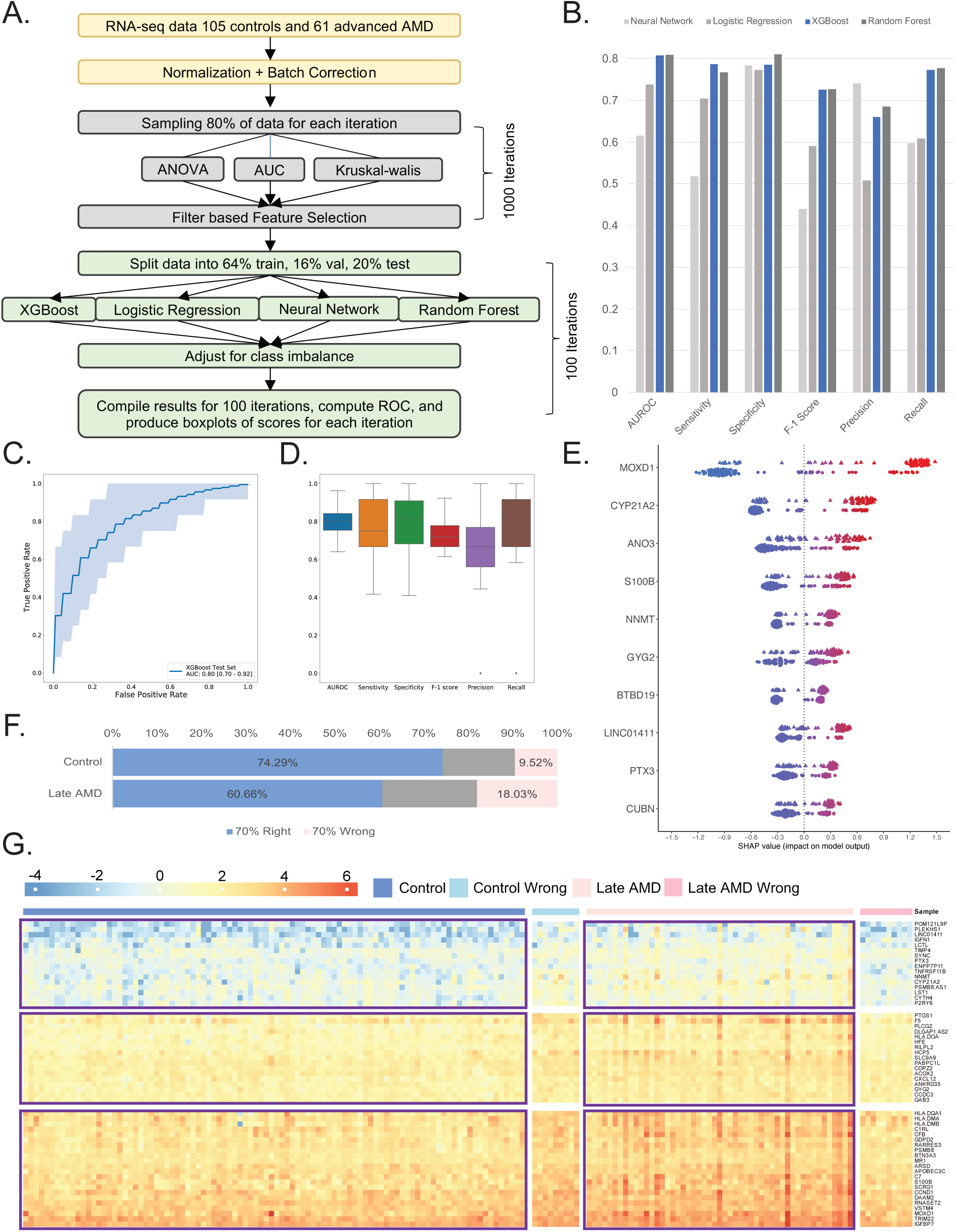
A flow-chart of ML-pipeline, models performance and results of late AMD classification. A. Schematic representation of Machine Learning pipeline, consisting of three main parts: normalization and batch correction, feature selection, and model building. B. Bar plot comparing each model’s statistics when used to classify between AMD cases and controls. Different colors represent the models built using logistic regression, random forest, neural networks, and XGBoost separately. C. An ROC plot showing the performance of the XGBoost model using default parameters. The closer the curve is to the top left corner, the more accurate the model will be at classifying cases and controls. The numbers presented at the bottom right represent the averages of 100 iterations of XGBoost models. D. Boxplot showing the distribution of statistics (AUC, Sensitivity, Specificity, F-1 score Precision and Recall) generated across the 100 iterations. E. Feature importance plot using SHAP analysis swarm plot showing the underlying weights of the top 10 genes for each sample. For each gene, the top swarm line shows the distribution of weights for AMD samples, while the bottom lines show the same information for Controls. The x-axis represents the SHAP value score each observation has within a gene. Observations are assigned colors corresponding to the range of gene expression, with dots (controls) and triangles (cases) closer to blue indicating lower gene expression values and those closer to red signifying higher gene expression values. F. Bar plot showing the distribution of samples being classified correctly 70% or more of the time, or samples being classified wrongly 70% or more of the time. The sections are identified as blue and pink respectively. Grey sections are for samples not making the right and wrong predictions cutoff. G. Heatmap showing the gene expression of 81 ML-genes (as rows) and 166 samples (as columns) divided into 4 groups: Controls, Controls being predicted wrong 70% or more of the time, Cases, Cases being predicted wrong 70% or more of the time.

Next, we applied four ML-based models: neural network, logistic regression, eXtreme Gradient Boosting (XGB), and random forest for the classification model for AMD based on 81 ML-genes. We randomly portioned the data into 64% training (to learn potential underlying patterns), 16% validation (to tune the model’s performance across different hyperparameter choices) and 20% (to evaluate our model’s prediction performance) external test sets. The optimal threshold for classification was determined by Youden’s J statistic^20^. We evaluated classifier training and discrimination performance in 100 iterations of repeated randomized data splitting to ensure the robustness of the model and obtain confidence intervals The AUC-ROC of other methods varied from 0.61 (CI 0.5-0.73) for Neural Network to 0.81 (CI 0.71-0.92) for random Forest (**Fig. 1B**) (**Supplementary Fig. 2**). XGB was found to perform the best with AUC-ROC statistic (0.80, CI 0.70-0.92) (**Fig. 1C**) and highest sensitivity (0.78) (**Fig. 1D**). Thus, we applied XGB for all further analyses.

To test the robustness of the 81 ML-genes identified in our study, we conducted comparisons of model performance using four additional gene lists: (1) Genes within 500KB of the 34 AMD-GWAS loci^9^ (2) High confidence AMD genes, comprising genes from the 34 loci with established connections to AMD through rare variant discovery or eQTL analysis (3) Genes deemed relevant to macular degeneration pathogenesis in the literature that emerged from extensive PubMed searched as previously described^13^ and (4) 48 genes identified through 1000 iterations of label shuffling, with control and AMD labels randomized. The performance of 81 features was superior compared to the genes within GWAS loci (AUC-ROC = 0.72, CI 0.58-0.84), high-confidence genes (AUC-ROC = 0.64, CI 0.50-0.77), literature (AUC-ROC = 0.69, CI 0.56-0.84) and, shuffled, i.e., permutation testing (AUC-ROC = 0.60, CI 0.45-0.75) (**Supplementary Fig. 3 A-D**). The 48 genes identified in the permutation testing showed no overlap with the set of 81 genes and performed poorly on both the true and shuffled labels (**Supplementary Fig. 4**). These results further emphasize the specificity of the 81 genes associated with AMD.

We next used SHAP (Shapley Additive exPlanations)^21^ to explain our best transcriptome-based AMD predictions by computing the contributions of each feature (gene) to that prediction (i.e., rank feature importance on classification). Shapley values indicate the contribution of every feature, i.e., gene expression value, towards the prediction for every individual sample, i.e., patient or control, vis-a-vis an average prediction. A positive Shapley value for a feature in each sample indicates that the feature value is favoring the prediction of the sample as the disease class with the magnitude of the Shapley value indicating the strength of how much it favors the prediction. A negative Shapley value for a feature in each sample can be considered vice versa. The results showed that high gene expression of *MOXD1* in AMD (red, triangle) and low gene expression of *MOXD1* in controls (blue, dots) contributed most to the model prediction. The trend was similar for the top 10 genes (**Fig. 1E)**.

Gene expression variation within humans arises from a complex interplay of genetic, environmental, and epigenetic factors. Furthermore, AMD manifests with a wide array of clinical presentations, encompassing both dry and wet forms, each exhibiting varying rates of disease progression and degrees of visual impairment. Thus, we next set out to identify whether such heterogeneity existed at the molecular (transcriptome) level. To achieve this, we harnessed the predictive capacity of the XGB model, training it on our dataset through 100 iterations of repeated randomized data splits. We compared the predicted labels against the actual ground truth (disease vs. control status) to uncover patterns. We systematically identified samples for which the predicted labels consistently aligned or deviated with the true labels in over 70% of instances. We categorize these samples into two groups: the "70% right" group, comprising instances where predictions align with true labels, and the "70% wrong" group, encompassing instances where predictions deviate from true labels. Notably, our analysis revealed a distinct pattern: a higher proportion of control samples (74%) exhibited accurate labeling compared to AMD samples (60%) (**Fig. 1F**). This discrepancy was further highlighted by the observation that nearly twice as many AMD patients were subject to mislabeling (18%) compared to only 9% of control subjects (**Fig. 1F**). Additionally, the performance of XGB was significantly improved (AUC-ROC = 0.94, CI 0.86-0.91) when 70% wrong samples were excluded from the analysis (**Supplementary Fig. 5**). This suggests that while heterogeneity exists within both groups, its manifestation is notably more pronounced within the disease population. These differences could not be attributed to age as within the same age range, there were several normal and AMD patients that were predicted accurately (**Supplementary Fig. 6A**). Next, we compared the top 2 risk alleles for AMD in *CFH* (Y402H; rs1061170) and *ARMS2* (A69S;rs10490924) as well as the polygenic risk scores (PRS) (based on 52 known common risk factors across 34 loci^9^ (**Supplementary Fig. 6C, D)**. We observed an expected, significant difference in CFH and ARMS2 risk alleles and PRS in all samples, and 70% right group. This difference was notably absent in the 70% wrong group (**Supplementary Fig. 6E**). These results suggest a potential involvement of genetic risk factors in shaping the molecular landscape of the disease in AMD. It’s important to note that the sample size remains small within the 70% wrong group (consisting of 10 controls and 11 AMD cases), underscoring the need for validation within larger cohorts. A heatmap of 81 ML-genes when plotted in these four groups (70% right AMD and controls, and 70% wrong AMD and controls) highlights the distinct gene expression patterns with the gene expression profiles within the 70% wrong group aligning closely with their predicted labels (**Fig. 1G**).

### Gene co-expression network-based analysis connect the ML-genes to AMD-relevant pathways

To gain further insight into the biological significance and relationships among the 81 genes, we utilized Weighted Gene Co-expression Network Analysis (WGCNA), known for its ability to associate gene co-expression modules with specific biological functions and pathways^22^. WGCNA analysis was done using transcriptome data from 453 human retina and identified 44 modules and used GO analysis to identify the top biological pathways associated with these modules. We observed that majority of the (62/81) ML-genes were enriched within three modules associated with immune response (turquoise, *p-value*= 2.07 x10^-6^), extracellular matrix organization (ECM) (tan, *p-value*= 5.16 x10^-19^), and complement (magenta, *p-value*= 3.45 x10^-19^) pathways (**Fig. 2A**). These results are particularly interesting because of the putative role for these pathways in the pathogenesis of^12, 23^. Additionally, these modules also harbor three known AMD-GWAS genes, *C3* and *COL8A1 (*tan) and *CFB* (magenta)^23^. *FBLN1*^24^ and *MOXD1*^13^ were particularly interesting for their implicated role in AMD. Additionally, we find several ML-genes involved in complement pathway such as *C7*, *C1S*, *C1R* and *C1RL* that have not been associated with AMD. Next, we assessed the module trait correlation across normal and AMD patients to identify the gene networks associated with the disease (**Supplementary Fig. 7**). Notably, all three ML-enriched modules exhibited a positive correlation with AMD and the eigengene for these modules demonstrated increased expression levels between normal and AMD (**Fig. 2B**).

**Figure 2:**
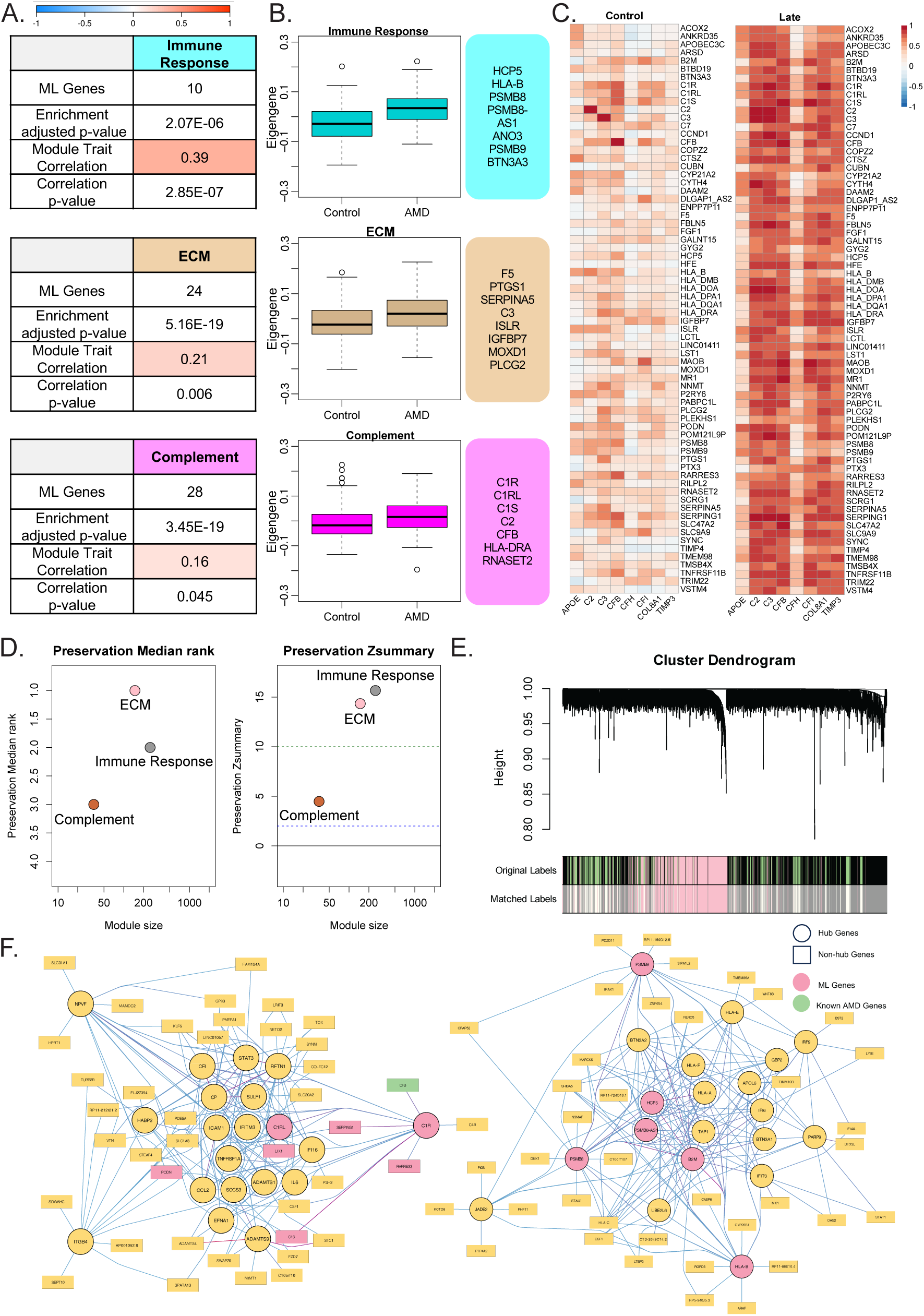
Gene co-expression network analysis to connect ML-genes to disease pathways. A. Table representing the number of ML genes included in the modules, the p-value of the enrichment, the correlation value between the module’s eigengene expression profile and the AMD status of the samples, and the correlation p-value. The color in each correlation cell corresponds to the correlation value on the scale provided at the top of the table. Notable, immune Response ranks highest in correlation to a patient being diagnosed with AMD. B. Boxplot showing the eigengene value of each module between cases and controls. Next to the boxplots are the subset of 81 ML-genes that were identified as part of the top 10 hub genes within those modules. C. Heat maps illustrating the correlation values between 81 ML-genes and known AMD-GWAS genes, categorized into control and AMD groups. ML-genes with a correlation value ≥0.7 with any known AMD genes in the AMD sample group are included. That list of genes was used to generate the heatmap for the Control group. D. Module Preservation plot showing the preservation of gene composition in immune response and ECM modules from the control network in the late AMD network. The module enriched for complement pathways was weakly preserved in the AMD network. E. Hierarchical cluster dendrogram of **control and AMD** co-expression networks. Each black branch (vertical line) corresponds to one gene. The color rows below the dendrogram indicate module membership showing the labels of genes in the Immune Response (Black), ECM (pink), and Complement (lightgreen) from the AMD network when matching with their labels in control network. F. A Cytoscape visualization of the top 20 hub genes and their top 10 connections to other genes. Round nodes are the top 20 hub genes while rectangular nodes depict connected genes. ML-genes are highlighted in pink, and known AMD-GWAS genes are highlighted in green. Connections between genes are color-coded, with purple indicating stronger connections, while blue represents weaker connections.

Expression correlations are often used to infer functionality and regulatory relationships within specific biological contexts. As we did not observe many known AMD-GWAS genes in 81 ML-genes, we further explored the functional relationship between known AMD-GWAS genes with the ML-genes identified in this study between controls and AMD patients’ transcriptome data. We observed a strong correlation (r2 >0.7) in cases compared to controls with eight known AMD-GWAS genes correlated with 70/81 ML-genes (**Fig. 2C**, **Supplementary Fig. 8**) in late AMD whereas in controls only 2 known AMD-GWAS genes were correlated with 7/81 ML-genes (**Supplementary Fig. 9**). Additionally, these correlations exhibited statistical differences between cases and controls for most ML-genes (**Supplementary Fig. 8**). In comparison, in random set of 81 genes, a much smaller number of genes showed correlation and they were comparable in cases (20/81) and controls (25/81) (**Supplementary Fig. 10).** Furthermore, and even fewer of these correlations were statistically different between cases and controls (**Supplementary Fig. 11**). Taken together, these findings demonstrate that there is a enhanced positive correlation and thus by extension functional relationship between the expression ML-genes with known AMD-GWAS genes in AMD transcriptomes.

Next, we analyzed the preservation of the three modules enriched for ML-genes in normal and AMD networks using the density and connectivity-based preservation statistics available within the modulePreservation in WGCNA. The overall measure of preservation was defined as Z_summary_ (**Fig. 2D, E**). The two modules that functionally annotate to immune response and ECM were well preserved between controls and AMD (Z^summary^ >10). However, the module enriched for complement pathway genes was found to be weakly preserved (Z_summary =_ 4.47)^25^. Next, we identified the top 20 most connected genes (hub genes) and their top 10 connections within the complement module from controls and AMD using WGCNA. We identified two ML-genes (*C1R* and *C1RL*) as hub genes in controls, whereas AMD network has six ML-genes (*PSMB8*, *PSMB9*, *PSMB8-AS1*, *B2M*, *HPC5*, *HLA-B*) as hub genes (**Fig. 2F**). These findings suggest these genes within complement pathways are modulators of immune activity in the retina that play an important role in pathogenesis in AMD.

### AMD disease progression has shared and unique gene signatures

AMD is a progressive disease with early, intermediate, and late stages of the disease. Early/intermediate AMD is the most common and asymptomatic form, characterized by pigmentary abnormalities in RPE of the macular region and accumulation of extracellular aggregates of proteins, lipids and cellular components (called drusen). Vision loss happens in the late stage, which is usually subdivided into dry (geographic atrophy, or GA) and wet (choroidal neovascularization, or CNV) forms^26^. The symptoms of AMD worsen over time, although the rate at which the disease progresses varies and not all patients with early/intermediate AMD develop late disease. In the United States alone, over 1.75 million people have late stages of AMD and 7.3 million people are affected with intermediate stages which are at the risk of developing late AMD^27^. However, there is a paucity of studies on the early and intermediate stages of AMD, and as a result, there are no reliable biomarkers for predicting the disease progression. Thus, we next applied the ML pipeline developed for late AMD in early (n= 175) and intermediate (n=112) AMD to identify the molecular events that lead to AMD. We identified a set of 57 genes for early AMD that provided AUC-ROC statistic of 0.62 (CI 0.51-0.74) (**Fig. 3A**), whereas a set of 62 genes gave AUC-ROC statistic of 0.71 (CI 0.59-0.83) for intermediate AMD **(Fig. 3B)**. The relatively modest performance of these models can be attributed to subtle alternations in gene expression during these initial stages, where vision loss or cell death in early and intermediate stages is not yet prominent. Thus, we next tested the performance of the features identified in early and intermediate AMD in late AMD. This analysis showed notable enhancement in predictive power with the 57 early AMD-associated genes, leading to an AUC-ROC statistic of 0.74 (CI 0.58-0.86) (**Fig. 3C**). Conversely, the performance remained comparable for the intermediate stage (AUC-ROC = 0.72, CI 0.58-0.89) **(Fig. 3D)**. For both stages, the performance of the features selected based on shuffled label did not perform well (**Supplementary Fig. 12**). These findings are also consistent with a lower sensitivity as well as a higher proportion of early (29%) (**Fig. 3E**) and intermediate AMD (27%) (**Fig. 3F**) deviating from their ground truth prediction. Thus, it is likely that intermediate AMD might have distinct molecular underpinning that does not represent a transitional stage between early and late AMD. This was also reflected in the expression correlation of the candidate genes with known AMD-GWAS genes. 81 ML-genes identified in the late AMD showed higher correlation in early AMD compared to the intermediate AMD (**Supplementary Fig. 13**). Similarly, early AMD 57 gene signatures also showed higher correlation with late AMD and not intermediate AMD (**Supplementary Fig. 14**). However, the gene identified in intermediate does not show correlation with known AMD genes in any stages **(Supplementary Fig. 15**). Importantly, genes identified across both early and intermediate stages were enriched within modules associated with immune response and ECM pathways (**Fig. 3G, H**). While there exists a limited overlap in genes identified among the three disease stages of AMD, they manifest enrichment within identical modules associated with AMD-relevant pathways. This underscores that majority of AMD-progression response includes signatures reflecting immune response dysregulation, indicating a shared biological basis.

**Figure 3:**
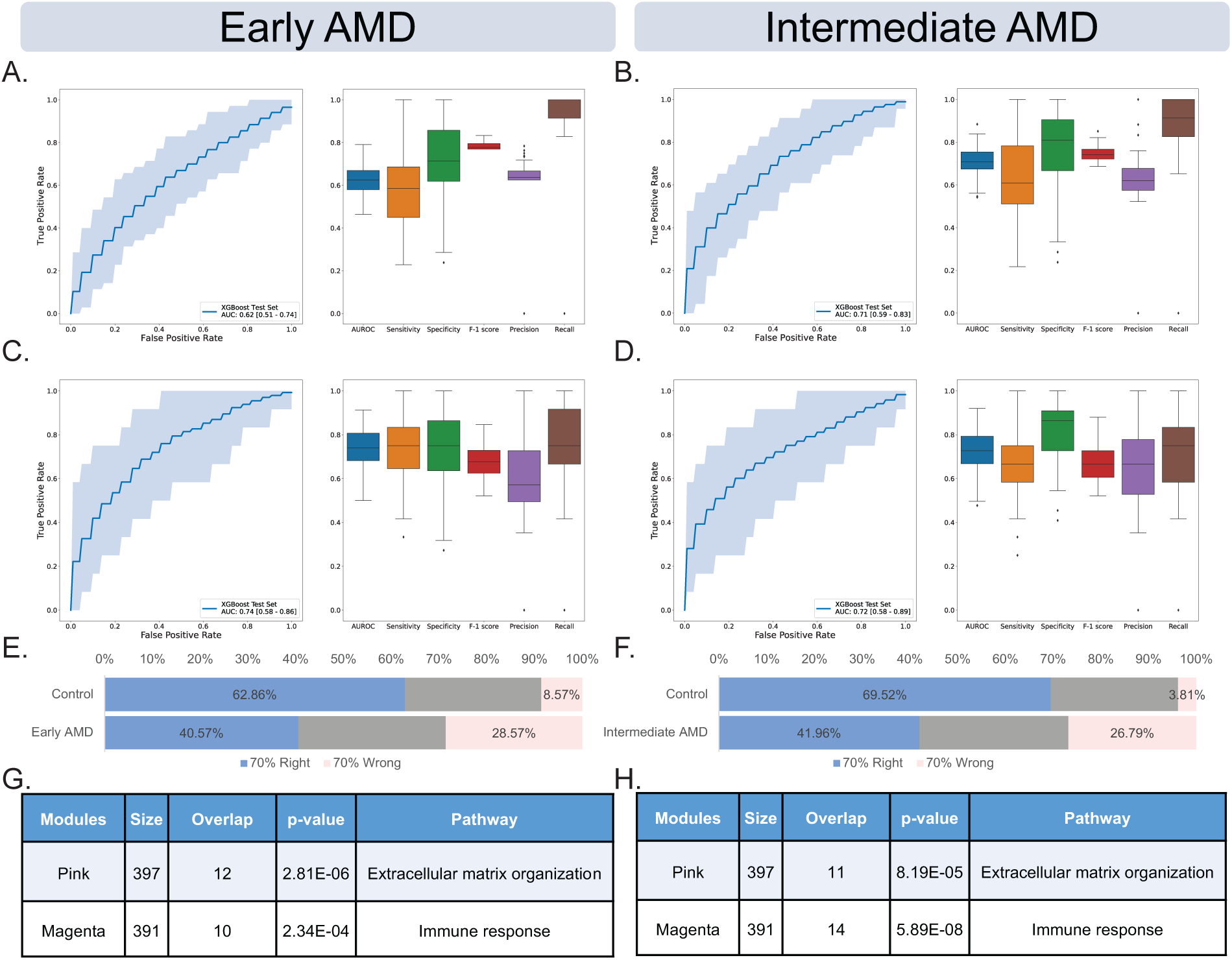
Performance ML methods in classifying early and intermediate-stage AMD. A. ROC and box plots illustrating the outcomes of classifying control and early AMD samples across 100 iterations using a set of 57 genes generated by the XGBoost model. B. ROC and box plots display the outcomes of classifying control and Intermediate AMD samples across 100 iterations using a set of 62 genes generated by the XGBoost model. C. ROC and box plots illustrate the notable enhancement in predicting late AMD outcomes by employing the XGBoost model across 100 iterations with 57 genes selected for Early AMD and control samples but applied to distinguish late AMD samples from the controls. D. ROC and box plots demonstrate that utilizing the XGBoost model across 100 iterations with 62 genes associated with intermediate AMD did not improve performance in predicting late AMD outcomes. E. A bar plot displays the distribution of early AMD and controls classified correctly 70% or more of the time (blue), or samples classified wrongly 70% or more of the time (pink). Grey sections represent samples that do not meet the criteria for either correct or wrong predictions. F. Bar plots distribution for the intermediate AMD. Only 40% of the intermediate AMD samples are predicted right compared to the ∼70% of the controls. G. Table demonstrating the enrichment of 57 early AMD genes within co-expression network modules associated with immune response and extracellular matrix (ECM) pathways, as determined by userListEnrichment within WGCNA. H. A table depicting the enrichment of the same pathways for the 62 intermediate AMD genes.

### ML-genes are expressed within specific cell types in retina that are impacted in AMD

Our RNA-seq data was performed at the tissue level and yielded an average of gene transcript abundance that reflects the average signal from mixtures of cell-type-specific gene expression levels. This is particularly relevant for tissues characterized by a highly heterogeneous cell type composition, such as the retina, which is made of six different cell types^28^. To understand the role of AMD-relevant cell type, we built a reference for the average expression of retinal cell types using cell-type specific markers^29^ from six human retinas across three different studies^30, 31, 32^ (**Supplementary Table 1**). A heatmap of 81 ML-genes across retinal cell types showed that the majority of them were enriched in their expression in microglia, astrocytes, müller glia and retinal ganglion cell (**Fig. 4A**). next, we implemented three distinct methods-CIBERSORTx^33^, dTangle^34^, and BayesPrism^35^—to deconvolute the cellular composition of both control and AMD samples. Subsequently, we applied a student t-test to identify the cell types exhibiting significant changes associated with the disease, revealing astrocyte, microglia, Müller glia, and rods proportion to be significantly different between normal and late AMD (**Supplementary Fig. 16**). Microglia, astrocyte, and müller glia proportion increase in the disease whereas the proportion of rods decreases (**Fig. 4B**). The decrease in the rods is observed only in the late stage, which could be the results of aging^36, 37^ as well as disease-related photoreceptor degeneration^38^. Notably, microglia were the only cell type that significantly changed in cell proportion across all stages of AMD, while alterations in astrocyte proportion were confined to early and late AMD stages. (**Fig. 4B**). The three tools used differ in their underlying algorithms, input requirements, and output formats. However, all of them point to the involvement of microglia in AMD, suggesting microglial activation and increased immune activity begin in early AMD much before the onset of photoreceptor loss in late AMD.

**Figure 4:**
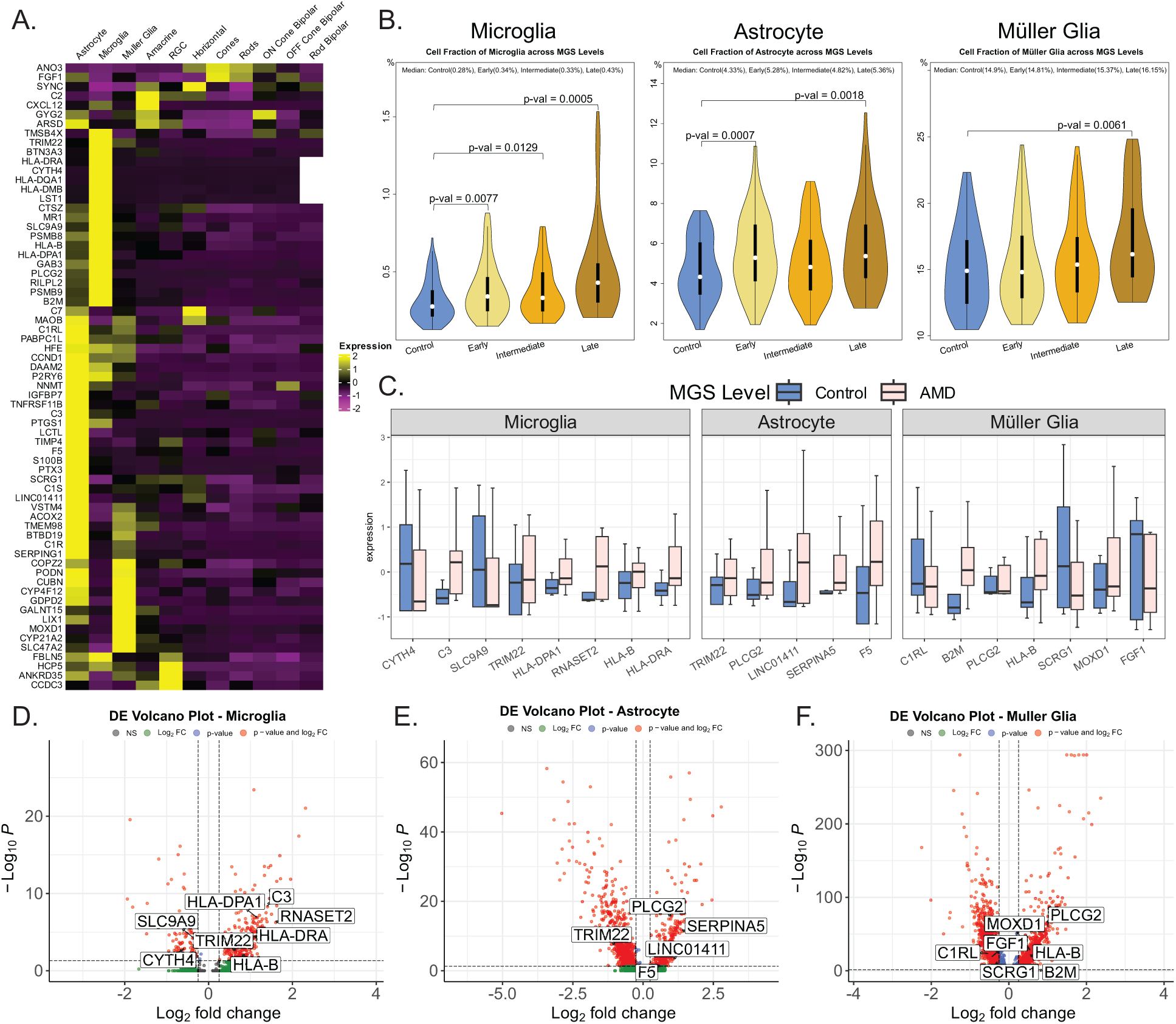
Expression of ML-genes across various retinal cell types and their alterations in AMD. A. A heatmap illustrating the average gene expression of 81 ML-genes (as rows) across 11 retinal cell types (as columns). The color gradient indicates whether genes are predominantly expressed (positive value, yellow) or minimally expressed (negative value, purple) in a particular cell type. Most ML-genes are expressed in astrocytes, microglia and Müller glia. B. Violin plots illustrating the cell fraction of various cell types in deconvolution results from 453 bulk RNA samples, utilizing a single-cell RNA dataset as a reference. P-values are annotated to indicate significant differences in cell fraction ranges between Control and different AMD stages. P-values are omitted when they exceed 0.05, indicating a lack of statistical significance. C. Box plots showing the differences in ML-gene expression between 13 normal and 16 AMD single nuclei data across astrocytes, microglia, and Müller glia. The selected genes are those that have successfully passed Differential Expression analysis utilizing DESeq2 with a false discovery rate (FDR) threshold of 5%. D. Volcano plot showing differentially expressed ML-genes within microglia, with 8 genes (6 upregulated, 2 downregulated) passing the DE threshold of fold change 1.5 or higher and FDR 5%. E. Volcano plot showing differentially expressed genes within astrocytes. 4 upregulated and 1 downregulated passing the DE threshold of fold change 1.5 or higher and FDR 5%. F. Volcano plot displaying significant ML-genes within Müller Glia (4 upregulated and 3 downregulated) using the same threshold as mentioned above.

To validate the results of the deconvolution, we analyzed single-nuclei data from 13 controls and 17 late AMD patients from two published studies^15, 39^ (**Supplementary Fig. 17**). We found several ML-genes (20/81) to be differentially expressed in microglia, astrocyte and Müller glia (**Fig. 4D**). Specifically, within the microglia population, we identified four subclusters. Notably, two of these subclusters, labeled as M1 and M2, displayed majority of the microglia cells and cell numbers between the AMD and control samples (**Supplementary Fig. 18**). Among these subclusters, a set of 8 genes were differentially expressed (6 upregulated, 2 downregulated (**Fig. 4D**). Within astrocytes, five gene were differentially expressed (4 upregulated and 1 downregulated) (**Fig. 4E**). In case of Müller glia, 4 genes were upregulated and 3 were downregulated (**Fig. 4F**). In addition, several known AMD genes including *APOE* and *VEGFA* were also found to be differentially expressed within glial population of normal and AMD patients (**Supplementary Table 2**). These findings collectively provide support for the consistency and validity of the genes identified using the ML approach and cell types identified using the deconvolution method in an independent dataset, reinforcing the relevance of the identified gene expression alterations in the context of AMD.

### AMD signature genes are enriched for AMD associated variants

Comparing transcript levels between healthy and diseased individuals cannot separate the cause vs consequences of the disease under scrutiny. Thus, we resorted to the published AMD-GWAS data on late AMD, comprising 16,144 patients and 17,832 controls^9^ as well as early AMD data consisting of 14,034 cases and 91,214 controls^40^ to access the potential association of genetic variants within ML-genes with AMD. The Quantile-Quantile (Q-Q) plot in late AMD-GWAS data (**Fig. 5A**) showed the largest deviation from the null p-value of the ML-genes identified in late AMD (red line) followed by early AMD (green line) suggesting that a subset of the ML-genes had genetic variants associated with AMD. In Early AMD data, the gene identified in early AMD showed the largest deviation (green line) succeeded by late AMD (**Fig. 5B**). Interestingly, neither dataset exhibited apparent deviation for intermediate AMD (indicated by the blue line) (**Fig. 5 A, B**). Furthermore, the ML-genes within the WGCNA modules enriched for complement and ECM organization individually also showed enrichment within late AMD-GWAS (**Fig. 5C**). By applying a suggestive association threshold (p-value <5 x10^-5^), we identified two candidates, *PLCG2*; rs4133124, p-value= 2.59 x10^-6^ (**Fig. 5D**) and *IGFBP7;* rs1718877, p-value= 2.83 x10^-6^ (**Supplementary Fig. 19A**) for late AMD and *USP7;* rs1471435, p-value= 7.27 x10^-6^ (**Supplementary Fig. 19B**) and *NEIL1*; rs11634109, p-value= 4.27 x10^-5^ (**Supplementary Fig. 19C**) for early AMD.

**Figure 5:**
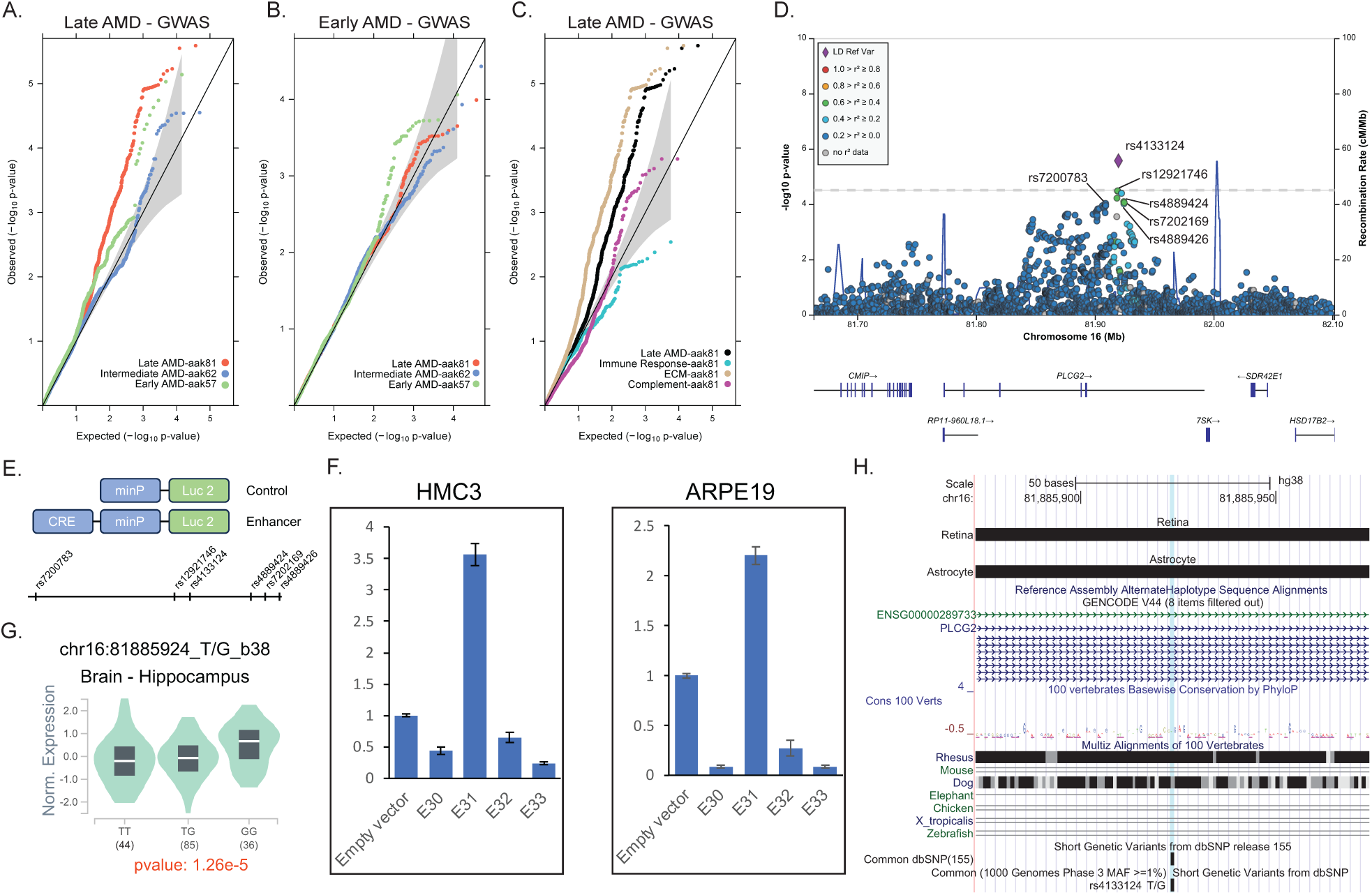
ML-genes are enriched for AMD-associated variants. A. Quantile-quantile (Q-Q) plot using summary statistics data from late AMD demonstrates a greater deviation from the null distribution (solid black line) for ML-genes identified in early (green dots) and late AMD (red dots) compared to intermediate AMD (blue dots). B. Q-Q plot using the early AMD data reveals similar but less pronounced trends across early, intermediate, and late AMD. C. Q-Q plot for the 81 ML-genes, further segregated into 3 groups based on the modules identified in the WGCNA network, also demonstrates deviation for the complement (purple dots) and ECM pathways (tan dots), but not for the immune response (turquoise dots). D. Regional association plot generated using LocusZoom plots displays the most strongly associated SNP, rs4133124 (purple diamond), along with other suggestively associated SNPs (p-value <5 x10^-5^) within the intron of PLCG2. E. A schematic representation of the luciferase assay, and the relative locations of the six SNPs around PLCG2 that were tested in the assays. F. eQTL violin plots sourced from GTEx to illustrate the correlation between the SNP rs4133124 and PLCG2 gene expression specifically within the hippocampus region of the brain. G. The luciferase assay results for four constructs (E30-E33) indicate that construct E31, which contains rs4133124, exhibits a 3.5-fold increase in luciferase activity compared to the empty vector in HMC3 and a 2.2-fold increase in ARPE19. Error bars represent the standard error of the mean (SEM) calculated from triplicate experiments. H. The UCSC Genome Browser graph displaying custom tracks from human retina, astrocytes, and ARPE19 cells shows the overlap of rs4133124 with open chromatin regions of AMD-relevant tissues and cell types. Additionally, the genomic region spanning rs4133124 shows conservation in primates as highlighted through multi-species alignment.

Next, we accessed the functional relevance of the suggestive associated SNPs, we selected four genomic regions spanning six SNPs around *PLCG2* and six genomics regions spanning seven SNPs around *IGFBP7* (**Supplementary Table 3**). We cloned these elements upstream of a minimal promoter-driven firefly luciferase gene in pGL4.23 (**Fig. 5E**) and tested for enhancer activity of the elements in the human microglia cell line, HMC3 and human RPE cell line, ARPE19. We identified that one element spanning rs4133124 within PLCG2 showed 3.5- and 2.2-fold higher luciferase activity compared to the empty vector in HMC3 and ARPE19, respectively (**Figure 5F**). We also tested the effect of reference T allele with the alternative G allele but found no change in enhancer activity in the rs4133124 region (data not shown). Additionally, this variant has been identified as an eQTL for *PLCG2* in hippocampus in the GTEx data(**Figure 5G)**^4^. We did not find this eQTL in the retina (data not shown), which could be attributed to small proportion of glial cell in the bulk retina data^13^. The variant, rs4133124 reside in the intronic region, which is highly conserved in primates, but not in mice (**Figure 5H)**. It is noteworthy that macular degeneration is also caused by the degeneration of photoreceptors and underlying RPE in the central region called macula, which is a primate specific structure^41^. Additionally, this variant resides within the open chromatin region in retina, ARPE19 and Astrocyte shown as custom track, suggesting a regulatory role (**Figure 5H)**. These results suggest that including the biological context of the genes can reveal additional genetic association within current GWAS datasets.

## DISCUSSION

For most complex diseases, including AMD, we have not exhausted the search for the disease genes as a significant proportion of heritability remains unexplained^9, 23^. Identification of additional loci warrants large case-control cohorts, which can be cost-prohibitive and limited by sample availability. Most AMD-GWAS variants reside in the non-coding region and mediate their effects through gene expression regulation^13, 15, 42^. Consequently, gene expression profiling in normal and disease samples provides valuable resource for studying disease mechanisms and discovering additional causal genes. Gene expression data exhibits high heterogeneity, with significant natural variation present within and among human populations^1^, a phenomenon exacerbated in diseases. Moreover, non-linear behavior is common in human systems due to their complex dynamics. Consequently, relying solely on a simple linear model, as often employed in the most common methods of differential gene expression analysis^43^, harbors inherent limitations and pitfalls. Additionally, arbitrary cutoffs of fold-change and statistical thresholds does not necessarily reflect biological relevance^44^. In contrast, our approach can detect and learn from non-linear data patterns to identify a robust molecular classifier through a series of rigorous feature recognition and dimensionality reduction.

Integration of prior knowledge from AMD biology with molecular networks be leveraged to understand the functional relevance of novel genes. The interconnected nature of gene regulatory networks implies that the expression of all genes in disease-relevant cells has the potential to influence the functions of core disease-related genes^45^. Co-expression networks are particularly useful for this purpose because when constructed using disease-relevant expression profiles, they can capture the tissue and cell-type-specific nature of the disease^46, 47^. It was notable that 76% (62/81) of ML-genes were involved in AMD-relevant immune response, complement and ECM pathways. Additionally, these genes had a much stronger pair-wise correlation with known AMD genes in cases compared to controls suggesting that during the disease process, they work closely with known AMD genes. Network preservation analysis further identified the modules involved in complement pathways to be not well preserved in the disease network affirming the well-established role of complement dysfunction in AMD^48^. The central regulatory hub genes of the complement modules were different in disease and controls. Among several ML-genes that were identified as hubs in the late AMD network, genes related to proteasome complex (*PSMB8*, *PSMB9*) are particularly interesting because of their role in immune system regulation^49^. It is therefore conceivable that dysregulation of proteasome activity-led immune response may contribute to the pathogenesis of AMD^50^. These results highlight the benefits of integrative analytical approaches to regain the holistic view of the AMD that is lost in experimentally tested reductionist approaches or hard statistical cut-offs.

Progression of early and intermediate to late AMD is observed frequently are attributed to multiple risk factors^51, 52^. However, the role of known genetic risk factors doesn’t seem to contribute significantly to the progression of intermediate to late AMD^53^. Additionally, in a sample of 6,657 cases of intermediate AMD, 10 out of 34 late AMD loci did not show association, despite having adequate statistical power^9^. This is further strengthened by our findings of intermediate AMD that showed modest improvement in model performance in late AMD. Similarly, digital deconvolution analyses revealed the changes in the astrocyte in the early and late AMD stage but not in the intermediate stage. However, the changes in the microglia proportion were observed across all three stages. Finally, integration of the AMD-GWAS data from early and late AMD does not show deviation for the genes associated with intermediate AMD. Taken together, our results suggest that AMD progression may not be linear and involve both shared and stage-specific genes, pathways, and cellular perturbations.

The dysregulated immune response is a hallmark of normal aging^54^ and a prominent feature in many neurodegenerative diseases^55^ including AMD^56^. However, molecular, and cellular mechanisms underlying immune dysregulation-mediated neurodegeneration are multifaceted and have not been completely resolved in AMD. In the human retina, immune responses are orchestrated by three distinct glial cell types: Müller cells, astrocytes, and microglia^57^. Microglia, akin to macrophages, serve as the resident immune cells and clear cellular debris through complement pathay^58^. They then maintain immune surveillance in the retina, supporting neuroprotection and homeostasis^58^ and can have different function based on anatomical location^59^. However, in disease conditions, they can get activated, migrate to the site of degeneration, and undergo morphological transformation leading to excessive release of inflammatory mediators and exacerbation of neurodegeneration^60^. Ocular sections from AMD samples shows the presence of activated microglia near the disease site that promotes degeneration^61^, and activated glial populations are enriched in AMD and related neurodegenerative diseases^39^. In contrast, a neuroprotective role of microglia has also been described in neurodegeneration^62, 63^. Our data further emphasizes the role of microglia at a molecular and cellular level in a large cohort (105 controls and 348 AMD). We show that genes associated with AMD have abundant expression in microglia and astrocytes. Secondly, we detected distinct differences in the cellular composition of microglia between normal and diseased individuals based on digital deconvolution of transcriptome profiles. These results suggest that gene relevant to AMD pathology modulate the retinal glial function that are driving force in disease progression and photoreceptor degeneration in AMD.

Reaching significant association signals (p<5×10^-8^) in traditional GWAS requires increasingly larger sample sizes to overcome statistical correction for multiple testing. Our approach of integrating the biological context, specifically genes exhibiting altered expression in disease-relevant cell type, with the GWAS data revealed novel genes within suggestive association signals (p<5×10^-5^). *PLCG2* encodes for an enzyme that catalyzes the hydrolysis of phospholipids and releases critical signaling messengers involved in diverse cellular functions^64^. Genetic variants in *PLCG2* have been associated with autoinflammation, antibody deficiency, and immune dysregulation syndrome^65^. Recently, the identification of *PLCG2* rare variants in Alzheimer’s patients has brought the focus on its role in neurodegenerative disease^66^, that is likely to cause the disease through microglia-mediated innate immune response^67^. A pathway-based analysis implicated the role of *PLCG2* in AMD^68^, however, our study presents the first convincing genetic and molecular evidence including the identification of rs4133124 as a single variant that is associated in both GWAS and eQTL analyses. The lack of observed differences in enhancer activity between the two alleles of rs4133124 could be due to the limitations of the luciferase construct used in the assay, potentially lacking crucial genomic elements necessary for detecting allele-specific effects. *IGFBP7* represents another such example which was identified as an AMD locus in the Japanese population^69^, but was not replicated in Caucasian-dominant AMD-GWAS^23^. Taken together, our study shows that the integration of gene expression data from normal and disease individuals with existing GWAS data provides a powerful approach for gaining systems-level insights into AMD pathogenesis.

## Materials and methods

### Cohort and data processing

We used RNA-seq data from 453 post-mortem donor retina that were evaluated to determine the level of AMD based on the Minnesota Grading System (MGS)^70^, with criteria similar to the Age-related Eye Disease Study (AREDS)^26^. MGS1 donor retina had no AMD features and served as controls, whereas MGS2-4 represented early, intermediate and late stages of AMD, respectively. Transcriptome analysis of donor retina was performed using RNA-Seq after enriching for poly-adenylated RNA. Raw RNA-Seq reads were processed as described earlier^13^. Briefly, trimmed reads were aligned to the Ensembl release 85 (GRCh38.p7) human genome using STAR version 2.5.2a^71^ RSEM^72^ was used to obtain estimated gene expression levels. Gene expression matrix was normalized using Trimmed Mean of M-values (TMM) in Counts per Million (CPM) using edgeR^73^ and genes were filtered by setting a threshold of 1 CPM in 10% of all samples. After initial quality control, 105 normal, 175 early, 112 intermediate and 61 late AMD samples were used in subsequent analyses.

### Feature selection

Normalized RNA-seq data was used to select the best features to be used in machine learning models. We applied three different feature selection methods ANOVA, AUC and Kruskal using mlr3filters (version 0.7.1) in R. All three methods are filter-based on scoring all the available features and then selecting features with the highest scores. ANOVA calculates the F-score to test the significance of each gene based on the analysis of variance. AUC computes a score called Area Under the Curve, also known as classification accuracy. The Kruskal method estimates the score for each gene utilizing the Kruskal-Wallis rank sum test, which is non-parametric compared to ANOVA.

We applied the feature selection 1000 times. For each iteration, we randomly sampled 80% of the data, used the three methods to obtain a score for each gene, and then filtered for the top five hundred genes. At the end of one thousand iterations, we obtained the list of genes and calculated how many times they were in the top five hundred in each iteration for each method. Finally, we took the top one hundred genes from each method and proceeded with the genes that appeared in all three methods.

### Machine learning models

We employed four machine learning models: neural network, logistic regression, eXtreme Gradient Boosting (XGB), and random forest. All machine learning models are available in Python as separate libraries. To train the model, we first randomly split the data into an 80% train and validation set and a 20% test set. Then we further split the train-validate set into eighty percent training set and twenty percent validation set. In other words, we randomly divided into 64% training set, 16% validation set, and 20% testing set. Subsequently, we fed the data into the models to train and generate predictions for each sample, validating the results. Once the models were trained, we evaluated their performance on the test set using metrics such as sensitivity, specificity, recall, precision, F-1 score, and AUROC. To handle the class imbalance between cases and controls, we applied the Youden J^20^ method to adjust the threshold for each iteration. The *p*-value cutoff was set to 0.05, and the best threshold was calculated using Youden’s J statistic^20, 74^. The maximum distance to the diagonal line was considered as the optimal cutoff point value. We then used the binary predictions to determine the frequency of correct predictions for each sample. Subsequently, we categorized the samples into two groups: those predicted correctly at least 70% of the time and those predicted incorrectly at least 70% of the time.

The following gene lists were used to compare the performance of chosen feature selection methods: **(1)** genes within 500KB of the 34 AMD-GWAS loci^9^ **(2)** High confidence AMD genes, including genes from the 34 loci for which the connection with AMD has been establish either through rare variant discovery, QTL analyses or functional validation **(3)** Genes deemed relevant to macular degeneration pathogenesis in the literature that emerged from extensive PubMed searched described before^13^ and **(4)** Features obtained by randomly swapping the labels of our dataset and rerun feature selection to obtain another set of genes.

We then tested both the original set of 81 genes and the later set of 48 genes obtained using shuffled labels by running the model using both sets. In one iteration, we ran the model with each set of genes using the true label. In another iteration, we shuffled all the labels in the training, validating, and testing set, and then ran the model. Finally, we ran the model with the training and validation set having shuffled labels, while keeping the labels of the testing set true.

SHAP (SHapley Additive exPlanations) was used for interpreting the output of the models by attributing the contribution of each feature to the final prediction. We used the model parameters built using the training data and the original gene expression as inputs for the SHAP library in Python to compute SHAP values for each instance^75^. The analysis shows a view of how each gene, or variable, will affect the model and alter the prediction.

### Weighted Gene-correlation Network Analysis

Weighted co-expression networks were constructed using the WGCNA^22^ using the Bioconductor R package. Briefly, a similarity matrix between each gene was obtained and the adjacency was calculated using Spearman correlation. We then used hypergeometric testing at a significance threshold of 0.05 alpha-level after Bonferroni correction accessing enrichment of genes for enrichment across identified modules. Pathway analysis was performed on each module using Gene Ontology biological process terms.

For module preservation, we first built the co-expression networks separately on controls and AMD samples. Next, we used the preservation statistics available within the modulePreservation function in the WGCNA^22^ in R. We then employed a permutation test (number of permutations = 500), which randomly permutes the module assignment in the control and AMD networks to assess if the observed value of preservation statistic is higher than what is expected by chance and assigns a permutation test *p*-value. The observed preservation values were then standardized with regard to the mean and variance and a significance Z score was defined for each preservation statistic. In order to compare the degree of preservation of modules between the normal and AMD networks, differential module preservation”—*ΔZ_summary_*, which is the arithmetic difference between the two preservation scores was calculated.

Hub genes were identified using the signedKME function, which calculated the KME values between a gene and all the modules in the network. With the corOptions parameter: “use = ‘p’, method = ‘spearman’”. This calculates the correlation between the expression patterns of each gene and the module eigengene. Genes with the largest kME are considered ‘hub’ genes within the modules. We selected the genes from their respective modules and sorted in descending order the KME value for that module. Genes with the largest kME we assigned ‘hub’ genes within the modules.

### Polygenic Risk Score

We obtained the beta coefficient of common, independently associated AMD-risk variants at 34 loci from published AMD-GWAS data^9^. 42 common, independently associated risk variants out of 52 were found in our data as the remaining were rare variants. To compute the polygenic risk score for each individual, we multiplied the genotype (coded as 0, 1, or 2) by its corresponding beta coefficient and sum up the weighted beta coefficient values across all variants. To test the difference between the PRS of Control and AMD, we used the Mann-Whitney U Test through the wilcox.test() function in R.

### Heat map

We created the gene expression heatmaps using pheatmap() function in R. First, we extracted the genes of interest’s expressions from the normalized bulk data and performed log 2-based transformation. The function pheatmap takes in a matrix. We defined the columns as the samples and the rows as the genes. Samples from control and AMD groups are separated to investigate the difference in gene expression between the two groups.

### Correlation with known AMD genes

We utilized the CPM normalized counts matrix to generate two distinct matrices: one comprising the 81 genes and the other containing the known AMD genes, separately for cases and controls. Subsequently, we employed the "cor" function in R to establish the correlation matrix between these two matrices. A threshold of 0.7 was applied, indicating that correlation coefficients greater than 0.7 or smaller than -0.7 would be considered significant. The "pheatmap" function in R was utilized to generate a correlation heat map between 81 ML-genes and known AMD genes between the control and AMD group. We used the “cocor” package in R to test the difference between the correlation of each gene pair in Control vs AMD. The “cocor” analysis was performed using a formula in the form "Gene1 + Gene2 | Gene1 + Gene2", where two independent datasets, the controls group, and the cases group were specified. cocor automatically selected Fisher’s test to determine the significance of differences between correlation coefficients. The resulting p-values were used to determine whether the correlation coefficients between the two groups were significantly different.

### Integration of GWAS data and Q-Q plot

We first removed of variants in the major histocompatibility complex region, and within +/- 1 Mb of the known GWAS signals^9, 40^. The data used for generating the Q-Q plot consists of a matrix derived from SNPs located within the gene bodies of the genes identified through feature selection (57 for early AMD, 62 for intermediate AMD and 81 genes for late AMD). Each entry in the matrix represents the negative base-10 logarithm of the quantiles for the corresponding SNP’s p-value. G-G Plot was used to plot the quantile on the x-axis and the minus log 10 p-value on the y-axis.

### Single Cell RNA-seq analysis

FASTQ files were downloaded from the GEO databases from three published studies: 2 samples from GSE202747^31^, 1 sample from GSE130636^32^ and 3 samples from the UK BioStudies database ^30^. Subsequently, sequencing reads were mapped to the available hg38 genome using CellRanger (version 6.1.2). The gene expression matrices generated by Cell Ranger were filtered to remove cells with unique molecular identifier (UMIs) less than 200 or more than 6000 or with more than twenty percent mitochondrial reads. Data was normalized and transformed using SCTransform from Seurat. To annotate the data with a UMAP visualization, we initially conducted dimensionality reduction using RunPCA and corrected batch effects with Harmony, followed by identifying nearest neighbors and clusters with FindNeighbors and FindClusters, and ultimately applying UMAP for visualization. Cells were annotated using a list of cell-type specific marker genes for 11 retinal cell types^29^. The DoHeatmap function within Seurat and used on SCT-normalized, scaled gene expression data to generate the heat map of the ML-genes within each cell type.

### Deconvolution

The bulk RNA-seq data containing 453 samples consisting of 105 control, 175 early AMD, 112 intermediate AMD, and 61 late AMD were used as mixture dataset^13^. We used Seurat-generated reference to implement three deconvolution methods: CIBERSORTx^33^, dTangle^34^, and BayesPrism^35^. For CIBERSORTx, the normalized count matrix of 3,956 differentially expressed genes across cell types was uploaded to the CIBERSORTx web application to generate the custom signature matrix of 2002. Normalized RNA-seq CPM counts data of the bulk data was used to perform the deconvolution. For dTangle on R, we used the CPM normalized counts RNA-seq counts data that was log2 normalized. The average expression of genes in the Seurat object was log2 normalized and used for dTangle. 14,705 genes that were present in both the bulk and single cell data were used in the deconvolution. BayesPrism analysis employed raw counts from our bulk RNA-seq dataset and SCT-normalized cell type-specific data from single-cell analysis. Genes on sex chromosomes and ribosomal genes were excluded, resulting in 13,716 genes used in the analysis. All three methods output cell fractions for each sample. Afterward, we performed a t-test on the cell fractions between the AMD samples and the control samples, specifically, samples from each stage of AMD against the samples from the control. From the cell fraction output of the deconvolution, we plotted the violin plots using “vioplot” package in R to visualize the difference in cell fractions across disease levels. We removed outliers in each disease stage level by setting a quantile range of 0.05 to 0.95.

### Single Nuclei Analysis

We used single nuclei data from human retina of 13 control and 17 AMD from two published datasets for this analysis^15, 39^. Counts data from 7 controls and 6 AMD patients were downloaded as raw FASTQ files were not provided^15^. FASTQ files from additional 6 controls and 11 AMD were downloaded from the GEO database (GSE221042)^39^ were processed using CellRanger (version 6.1.2). Data was analyzed within SEURAT as described above. Briefly, QC for each dataset was performed separately. We then performed SCTransform V2 on the merged Seurat object and then ran Harmony grouping by dataset and sample IDs. Next, we used FindNeighbor, FindClusters, and RunUMAP to cluster the cells and annotated the clusters using the list of cell-type specific marker genes. We then extracted the microglia, astrocyte, and müller glia clusters out of the Seurat object and redid the clustering to identify subclusters within each cell type. From there, we performed Differential Expression using FindMarkers (logfc_threshold = 0.1, min.pct = 0.1, test.used = ‘wilcox’) to identify differentially expressed genes between controls and AMD. Volcano plots were generated using “EnhancedVolcano” R package.

### Luciferase assay

We cloned 10 genomic regions spanning 13 SNPs (6 in PLCG2, and 7 in IGFBP7) upstream of a minimal promoter-driven firefly luciferase gene in pGL4.23 (Promega). HMC3, and ARPE19 cells were plated in 24-well plates (28K cells/well) and were transiently transfected after 24 hours with test luciferase constructs (500 ng) and *Renilla* luciferase vector (10 ng for transfection normalization) in triplicate using 2 μL of FuGENE HD transfection reagent (Roche Diagnostic) in 100 μL of Opti-MEM medium (Invitrogen). Cells were grown for 48 h and luminescence was measured using a dual luciferase reporter assay system on a Texan Spark Multimode Microplate Reader per the manufacturer’s instructions.

## Data Availability

The transcriptome data from 453 human donor retina used in this study are available in GEO (accession code <u>GSE115828</u>). Summary Statistics of advanced AMD is available at http://amdgenetics.org/ and early AMD is available for the download from www.genepi-regensburg.de/earlyamd. Single-cell retina data are available from <u>GSE202747</u> and <u>GSE130636</u> and single nuclei data was available under the accession code <u>GSE221042</u>.

## Code availability

All code used in this study was previously published and no customized code was used in this manuscript.

## Acknowledgements

This study is supported by Career Development Award from Research to Prevent Blindness (RPB) and New Investigator Award by BrightFocus Foundation to RR. RR is also supported by an Unrestricted grant from Research to Prevent Blindness (RPB) to Baylor College of Medicine.

## Author contributions

R.R. conceived and designed the study. K.M., H.N. A.B. and R.R. conducted the computational analysis and analyzed the data. N.B. performed the luciferase assays. K.M. and R.R. wrote the manuscript. All authors were involved in manuscript revision.

## Ethics declarations

The other authors declare no competing interests.

