## Supplementary file for "Integrating explainable machine learning and transcriptomics data reveals cell-type specific immune signatures underlying macular degeneration"

**Supplementary Information**

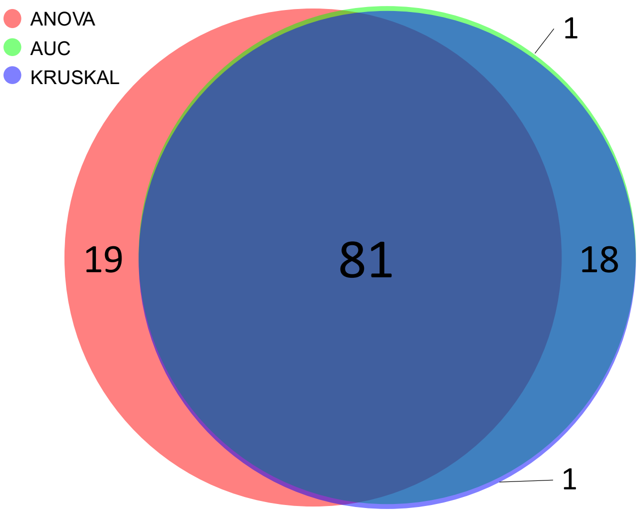

**Supplementary Figure 1**

Venn diagram illustrating the overlap between gene lists outputed by ANOVA, AUC, and Kruskal filtering models.

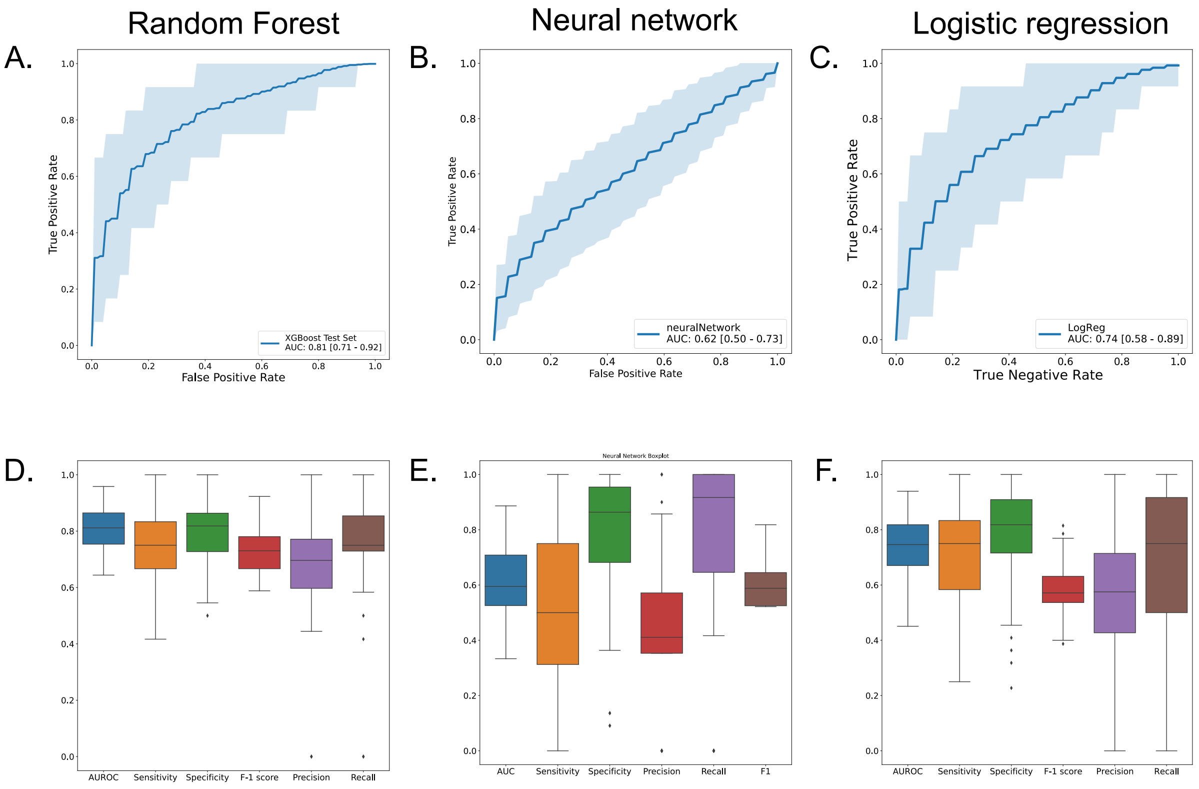

**Supplementary Figure 2**

(A-C) ROC plots showing the performance of ML-genes in Random Forest **(A)**, Neural network **(B)**, Logistic regression **(C)** using default parameters across 100 iterations.

(D-F) Boxplots showing different metrics Random Forest **(D)**, Neural network **(E)**, Logistic regression **(F)** across the 100 iterations.

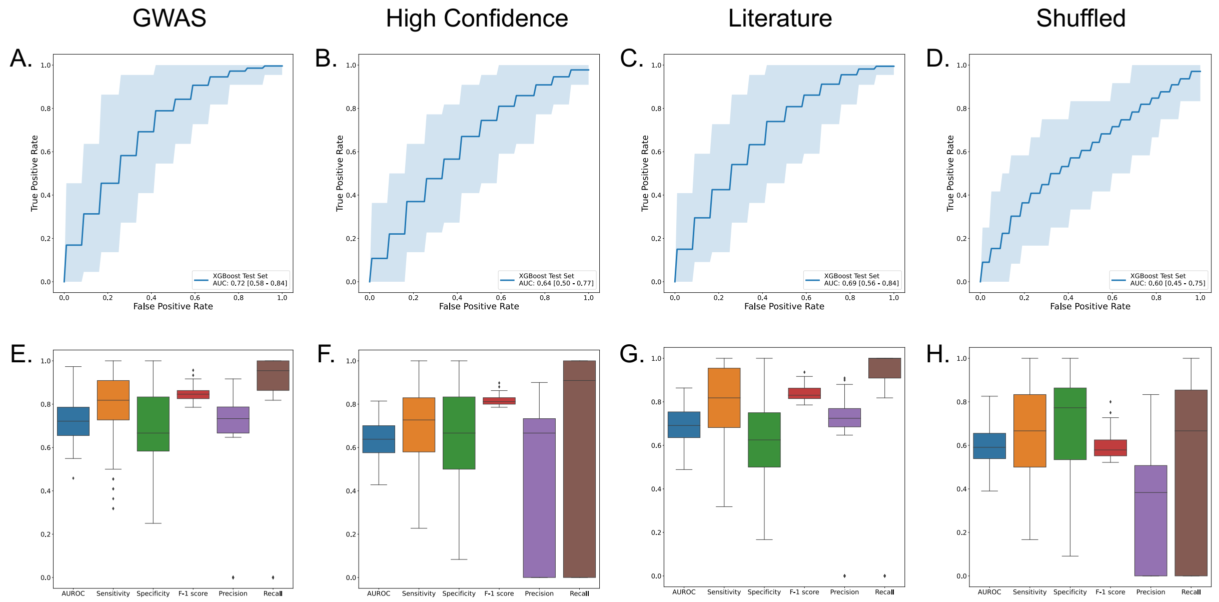

### Supplementary Figure 3

(A-D) ROC plots showing model performance on different subsets of training data randomly selected 100 times across different gene lists using XGBoost.

(E-H) Boxplots showing different metrics of measuring performance, including AUROC, Sensitivity, Specificity, F-1 score, Precision, and Recall for the 4 XGBoost models trained using different gene lists.

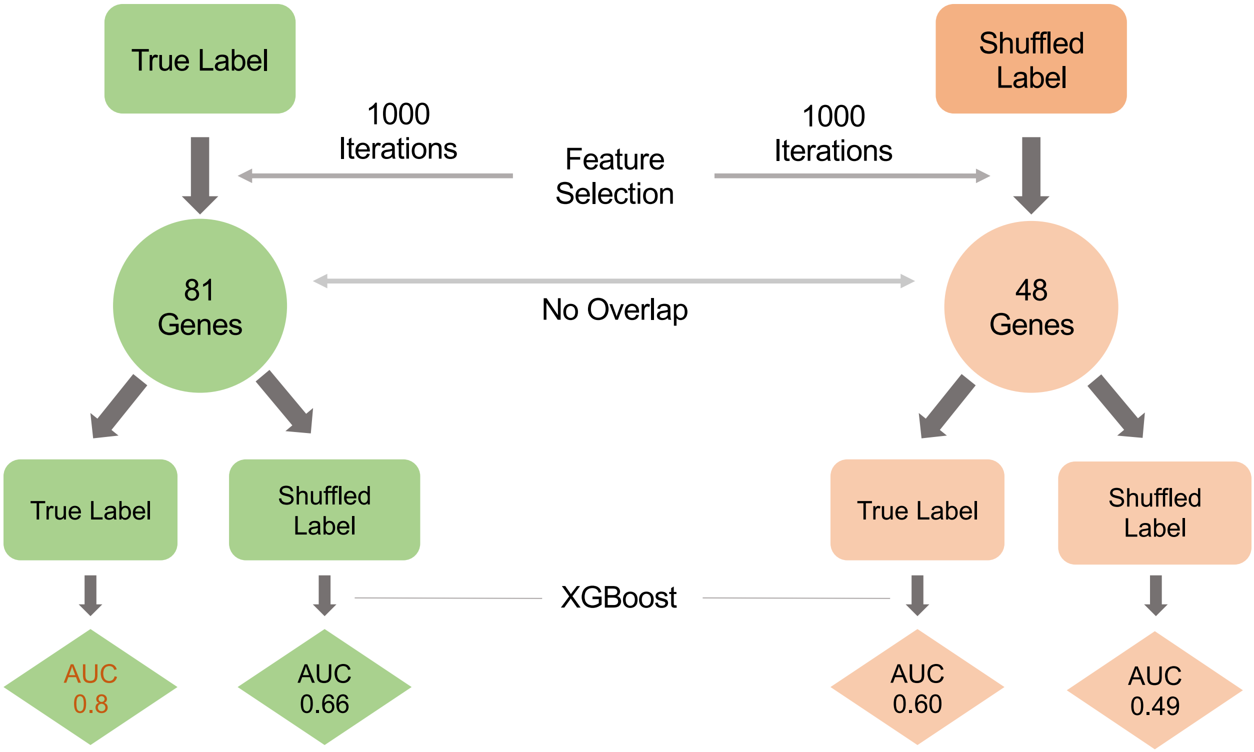

**Supplementary Figure 4**

Flow chart showing the process of training different XGBoost models using data with false labels to test the specificity of the genes selected.

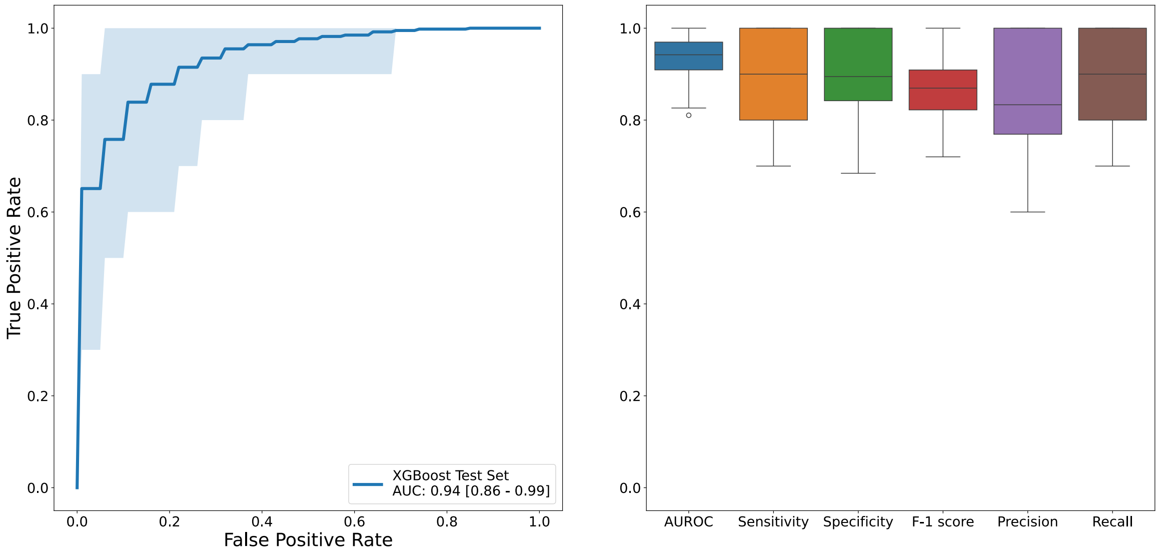

### Supplementary Figure 5

ROC plots showing model performance in 100 iterations using subsets of training data by removing “70% Wrong” samples.

A.

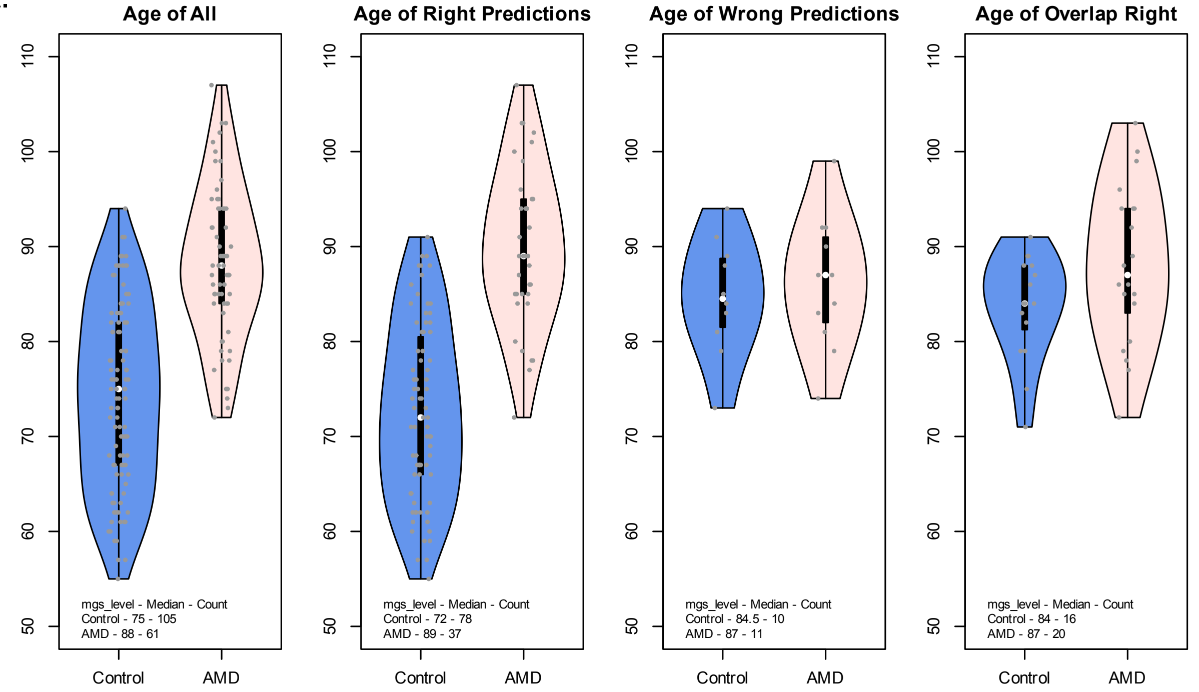

B.

| Test | All | Right Predictions | Wrong Predictions | Trimmed Right Predictions |
|---|---|---|---|---|
| Mann Whitney | 9.12E-15 | 6.03E-13 | 0.698 | 0.1107 |

C.

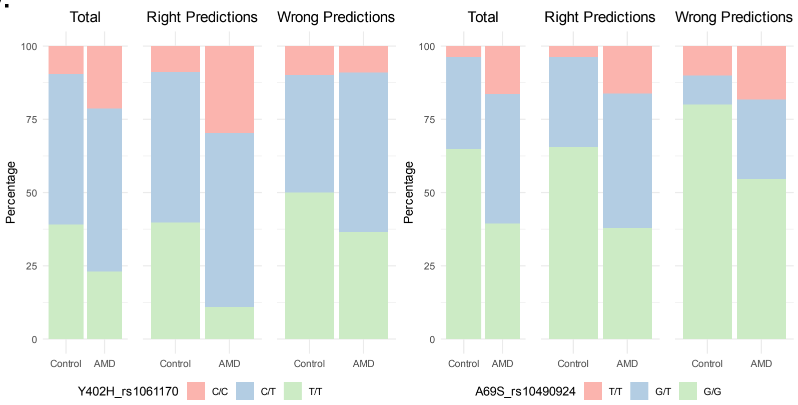

D.

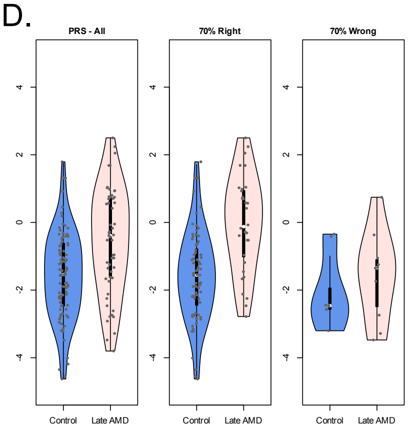

E.

| Y402H rs1061170 | ChiSq pvalue |
|---|---|
| Total | 0.001383 |
| Right | 2.22E-06 |
| Wrong | 0.6065 |

| A69S rs10490924 | ChiSq pvalue |
|---|---|
| Total | 3.01E-08 |
| Right | 1.86E-05 |
| Wrong | 0.0859 |

F.

| PRS | Mann Whitney |
|---|---|
| Total | 9.926E-05 |
| Right | 3.644E-06 |
| Wrong | 0.6334 |

**Supplementary Figure 6**

(A) Violin plots showing the distribution of age in all samples (n = 166), samples that were predicted right 70% of the time or more (n = 115), samples that were predicted wrong 70% of the time or more (n = 21), samples from the 70% Right group that have similar age range to the wrong group (n = 36)
(B) Table containing p-values of Mann Whitney, Kolmogorov-Smirnov shows that the distribution of data samples between Control and AMD groups were not significantly different in the right or wrong group.
(C) Stacked bar plots showing the distribution of the genotyping for two risk variants, A69S_rs10490924 and Y402H_rs1061170 across samples in the first three groups mentioned in (A). Allele associated with disease is at the top of the stacked bar plots.
(D) Violin plot showing the distribution of Polygenic Risk Score (PRS) calculated using the beta value from the AMD-GWAS data. The calculation was performed on samples that were genotyped (n = 406), utilizing the beta values of 44 variants across 34 loci. These samples were subsequently divided into groups as outlined in (A).
(E) Tables containing p-values from Chi-Square test and Proportion Z-test to test whether the proportion of the two risk variants are significantly different between Control and AMD. While the risk allele distribution was significantly different between cases and control in the 70% right group, the wrong group did not show any difference.
(F) Tables containing p-values from Mann Whitney and Kolmogorov-Smirnov significance of the difference between the PRS of Control and AMD in the 70% right group, which was notably absent from the 70% wrong group.

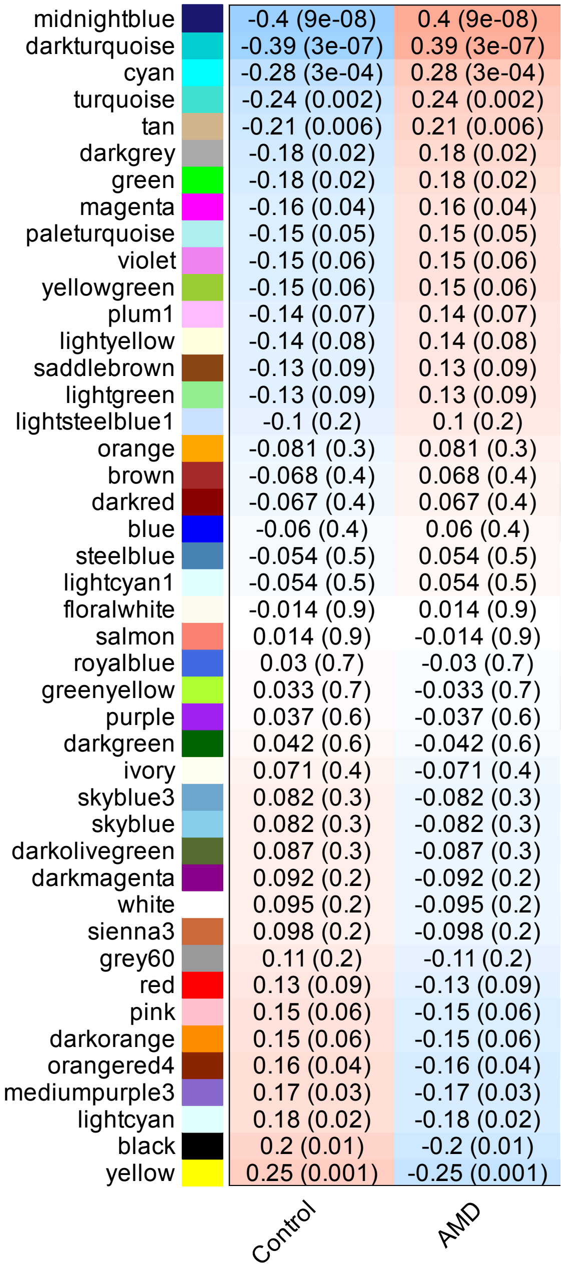

**Supplementary Figure 7**

Heatmap showing the correlation between the Eigengenes modules (using colors in R as temporary names) expression profiles against Control and AMD patients.

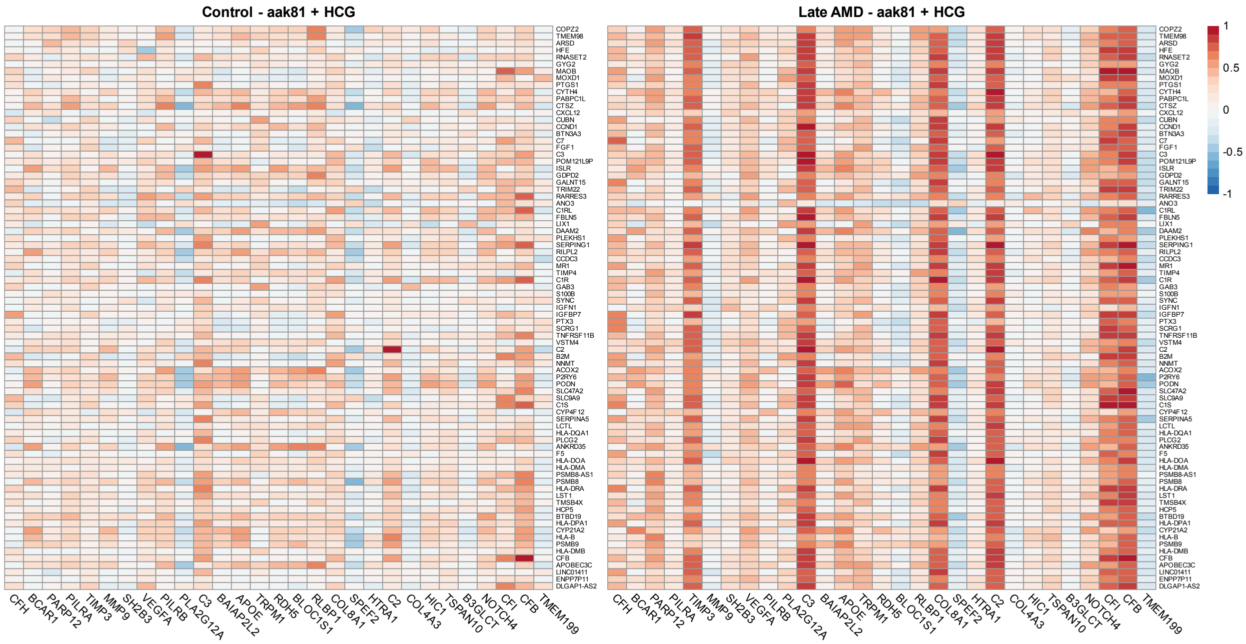

| AAK81 | Control | Late |
|---|---|---|
| ML genes | 7 | 70 |
| HC genes | 2 | 8 |

**Supplementary Figure 8**

Heatmap showing the difference in correlation between the 81 ML-genes and the known AMD genes (High Confidence Genes, HCG) based on the expression in Control and AMD samples. The table below contains the number of genes having a high correlation of 0.7 or more in the two groups.

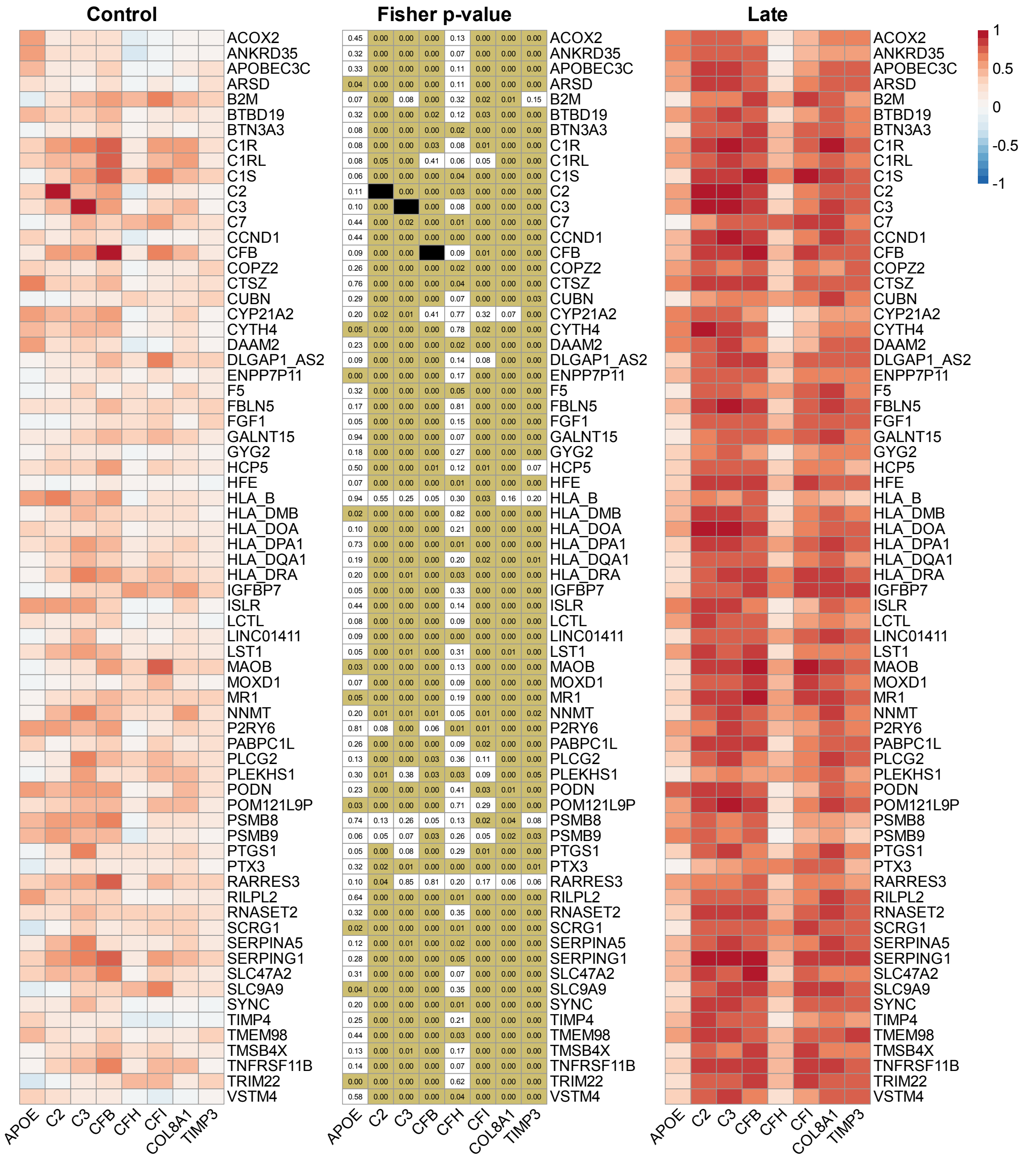

### Supplementary Figure 9

Heatmap showing the correlation between the 81 ML-genes list and the known AMD genes that passed the correlation greater than 0.7 in late AMD. The added middle panel shows the fisher p-value calculated using *CoCoR* package in R to test the significance of the difference between the correlation of each gene pair in Control versus AMD samples. Black cells in the center panel are due to the test being performed on the gene’s correlation with itself, resulting in null values.

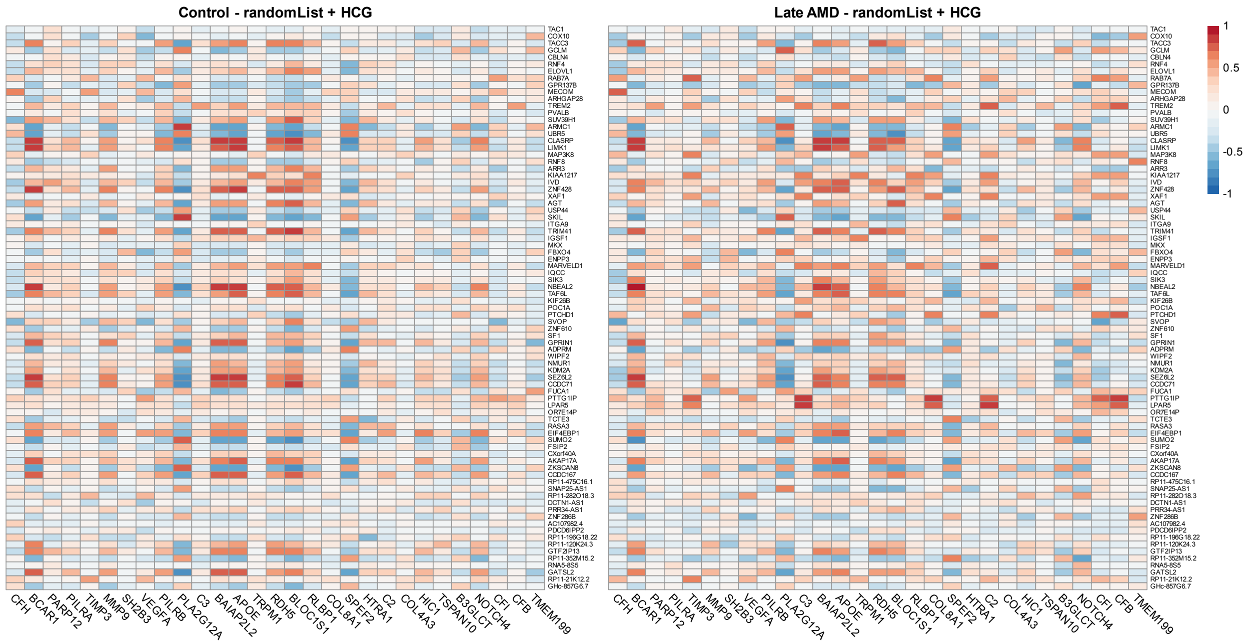

| Random List | control | late |
|---|---|---|
| ML genes | 25 | 20 |
| HC genes | 10 | 15 |

**Supplementary Figure 10**

Heatmap showing the correlation between the gene expression of random list of 81 ML-genes against the expression of known AMD genes. The table below contains the number of genes having a high correlation of 0.7 or more in the two groups were almost similar.

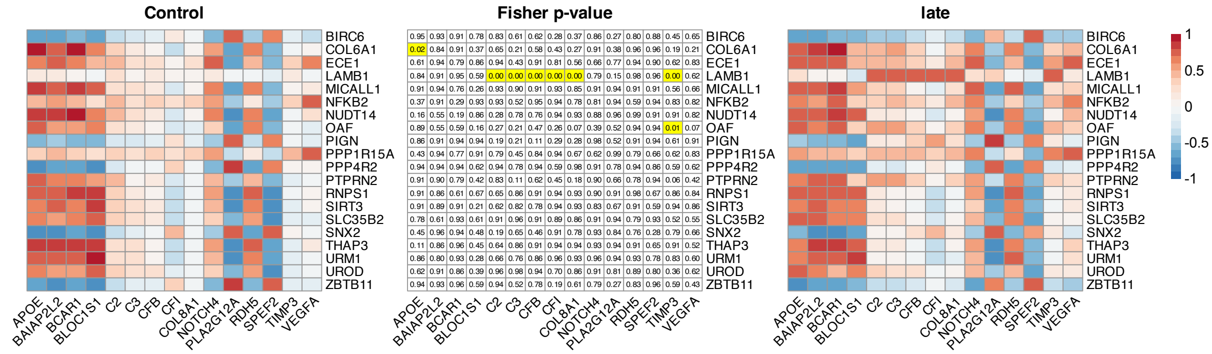

**Supplementary Figure 11**

Heatmap made using *ComplexHeatmap* library in R to show the correlation between a random list of 81 ML-genes and the known AMD genes, narrowed down to the ones passing 0.7 in correlation in late AMD. The middle panel shows the Fisher p-value calculated using CoCoR package in R to test the significance of the difference between the correlation of each gene pair in Control versus AMD samples.

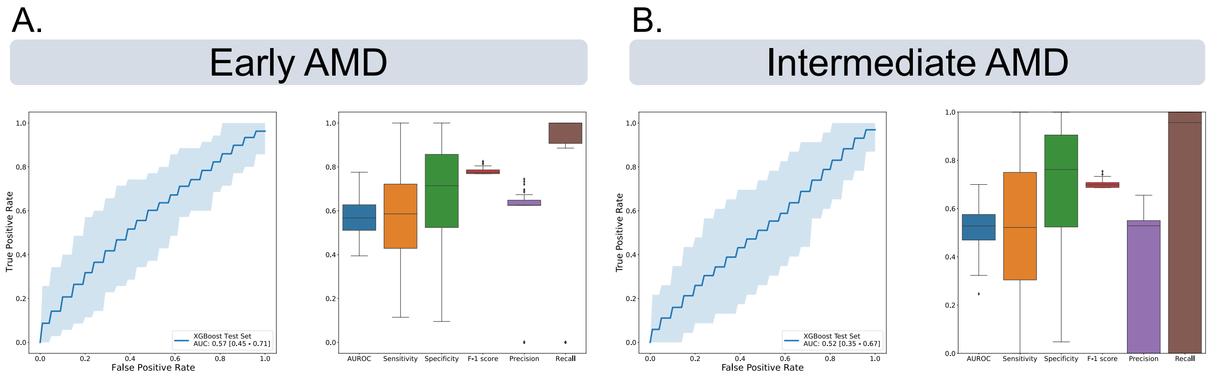

**Supplementary Figure 12**

A. The ROC curve plot and model statistics to show the performance of the XGBoost model of genes selected based on shuffled labels of Early AMD versus Controls.
B. The ROC curve plot and model statistics to show the performance of the XGBoost model of genes selected based on shuffled labels of intermediate AMD versus Controls.

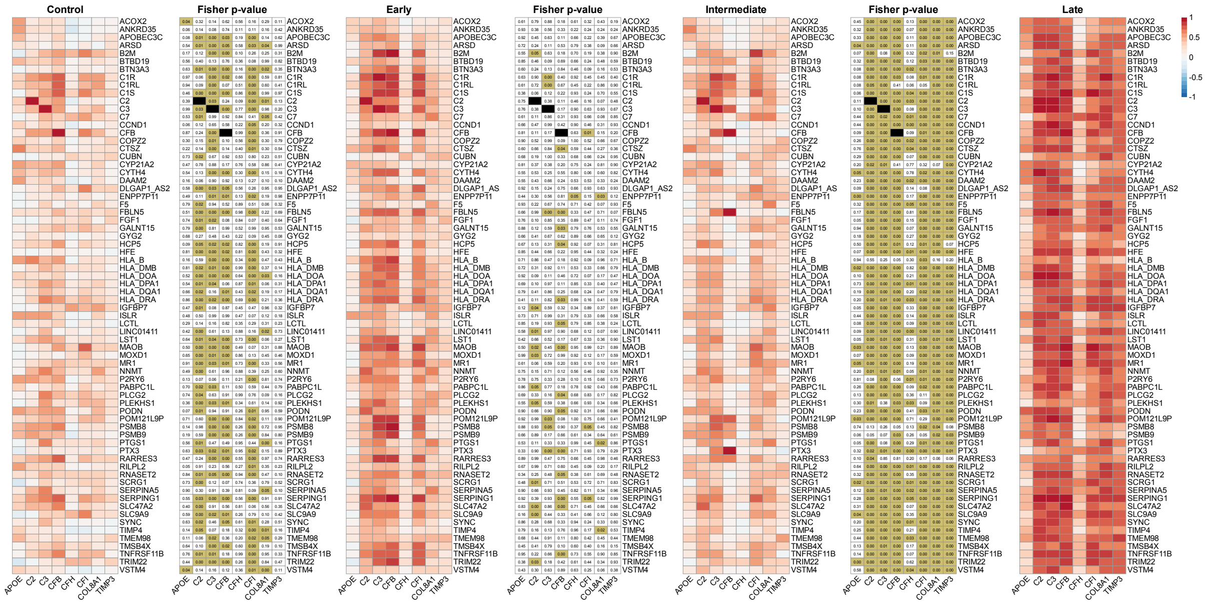

### Supplementary Figure 13

Heatmaps showing the difference in correlation values between 81 ML-genes and known AMD genes along with the Fisher's test p-value that shows the significant difference across all AMD stages when compared to the controls. The set of genes was obtained when selecting the genes having a correlation value ≥ 0.7 with at least one of the known AMD genes in late AMD samples.

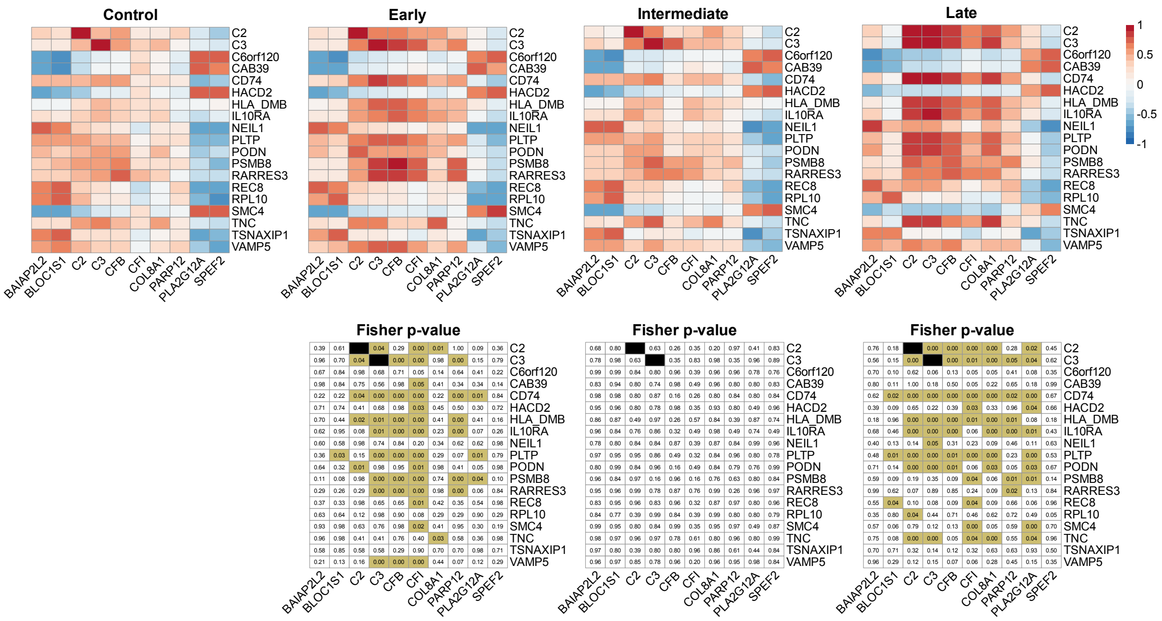

### Supplementary Figure 14

Heatmaps showing the difference in correlation values between 57 and known AMD genes along with the Fisher's test p-value to confirm the significant difference when comparing all AMD stages against Control samples. The set of genes was obtained when selecting the genes having a correlation value ≥ 0.7 with at least one of the known AMD genes in Early AMD samples.

**A**

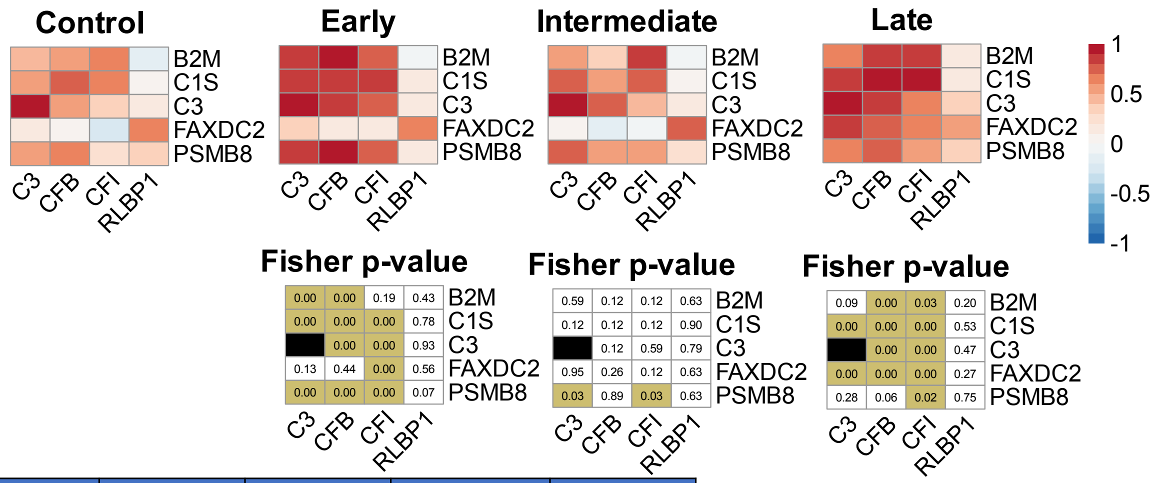

**B**

| aak57 | control | early | intermediate | late |
|---|---|---|---|---|
| ML genes | 14 | 19 | 19 | 34 |
| HC genes | 7 | 10 | 9 | 11 |

| aak62 | control | early | intermediate | late |
|---|---|---|---|---|
| ML genes | 1 | 12 | 5 | 43 |
| HC genes | 1 | 5 | 4 | 7 |

| aak81 | control | early | intermediate | late |
|---|---|---|---|---|
| ML genes | 7 | 34 | 16 | 70 |
| HC genes | 2 | 5 | 5 | 8 |

**Supplementary Figure 15**

(A) Heatmaps showing the difference in correlation values between 62 genes and known AMD genes along with the Fisher's test p-value to confirm the significant difference when comparing all AMD stages against Control samples. The set of genes was obtained when selecting the genes having a correlation value ≥ 0.7 with at least one of the known AMD genes in Intermediate AMD samples.

(B) Table showing the number of ML genes having a high correlation (≥ 0.7) with known AMD genes (HC genes) based on the heatmaps in Supplementary Figures 13,14, and 15.

A.

| CIBERSORTx | | | |
|---|---|---|---|
| Cell Type | Early | Intermediate | Late |
| Amacrine | 0.3420 | 0.9808 | 0.5234 |
| Astrocyte | 0.0022 | 0.0952 | 0.0019 |
| Cones | 0.3530 | 0.9790 | 0.7404 |
| Horizontal | 0.6921 | 0.2973 | 0.1389 |
| Microglia | 0.0007 | 0.0233 | 0.0004 |
| Muller Glia | 0.2407 | 0.1012 | 0.0058 |
| OFF Cone Bipolar | 0.6536 | 0.1376 | 0.0278 |
| ON Cone Bipolar | 0.3743 | 0.7654 | 0.3915 |
| RGC | 0.0530 | 0.8679 | 0.8911 |
| Rod Bipolar | 0.2809 | 0.9584 | 0.4128 |
| Rods | 0.1486 | 0.0966 | 0.0038 |

| dTangle | | | |
|---|---|---|---|
| Cell Type | Early | Intermediate | Late |
| Amacrine | 0.6391 | 0.4095 | 0.8799 |
| Astrocyte | 0.0062 | 0.0751 | 0.0011 |
| Cones | 0.0999 | 0.0535 | 0.0049 |
| Horizontal | 0.6478 | 0.2301 | 0.177 |
| Microglia | 0.0006 | 0.0256 | 0.0003 |
| Muller Glia | 0.3360 | 0.1295 | 0.0104 |
| OFF Cone Bipolar | 0.8338 | 0.2566 | 0.306 |
| ON Cone Bipolar | 0.8867 | 0.4798 | 0.2254 |
| RGC | 0.3421 | 0.4693 | 0.07619 |
| Rod Bipolar | 0.9483 | 0.8063 | 0.3593 |
| Rods | 0.1820 | 0.1338 | 0.0134 |

| Bayes Prism | | | |
|---|---|---|---|
| Cell Type | Early | Intermediate | Late |
| Amacrine | 0.5208 | 0.6072 | 0.9255 |
| Astrocyte | 0.0007 | 0.1013 | 0.0018 |
| Cones | 0.4903 | 0.2025 | 0.1785 |
| Horizontal | 0.5698 | 0.1227 | 0.0470 |
| Microglia | 0.0077 | 0.0129 | 0.0005 |
| Muller Glia | 0.4598 | 0.1896 | 0.0062 |
| OFF Cone Bipolar | 0.3443 | 0.08124 | 0.131 |
| ON Cone Bipolar | 0.21 | 0.5911 | 0.8405 |
| RGC | 0.5386 | 0.3811 | 0.3544 |
| Rod Bipolar | 0.2212 | 0.4624 | 0.7675 |
| Rods | 0.1793 | 0.1285 | 0.0117 |

B.

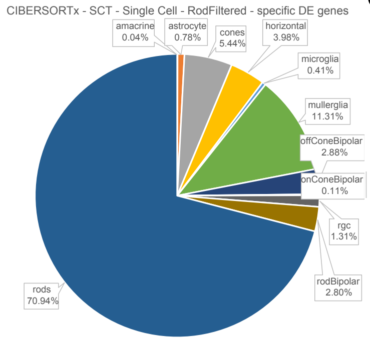

C.

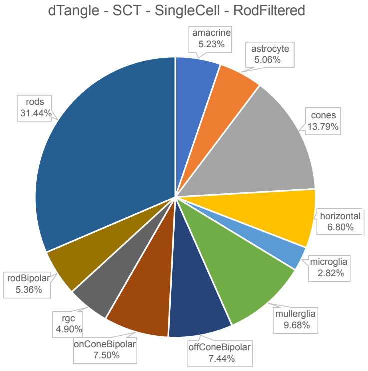

D.

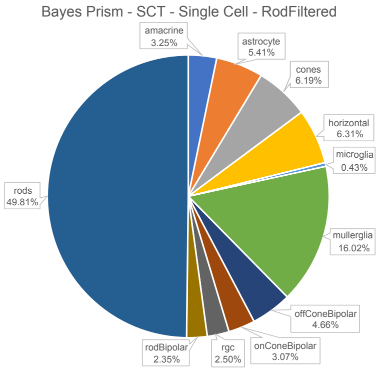

**Supplementary Figure 16**

(A) T-test table showing p-values of testing the difference between the fraction of cells after deconvolution between different stages of AMD compared to the control.
(B) Pie chart showing the average fraction of cells after deconvolution using CIBERSORTx.
(C) Pie chart showing the average fraction of cells after deconvolution using dTangle.
(D) Pie chart showing the average fraction of cells after deconvolution using Bayes Prism.

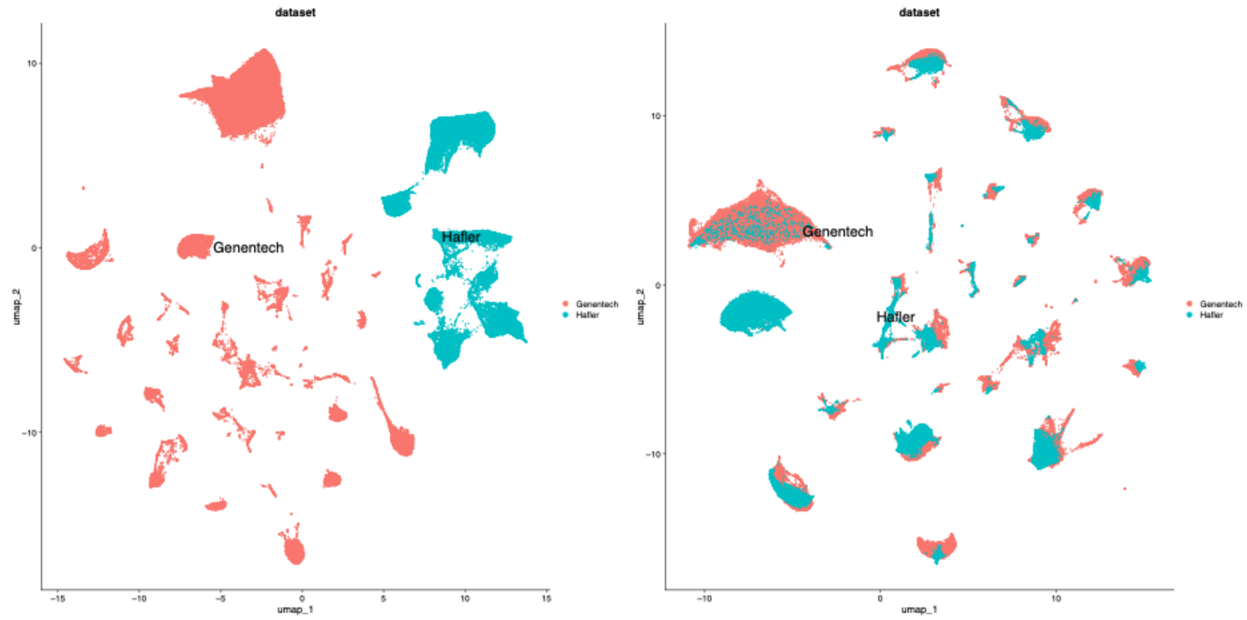

**Supplementary Figure 17**

UMAP showing the projected cell clusters between two datasets before and after using *RunHarmony* in R to eliminate batch effects.

A.

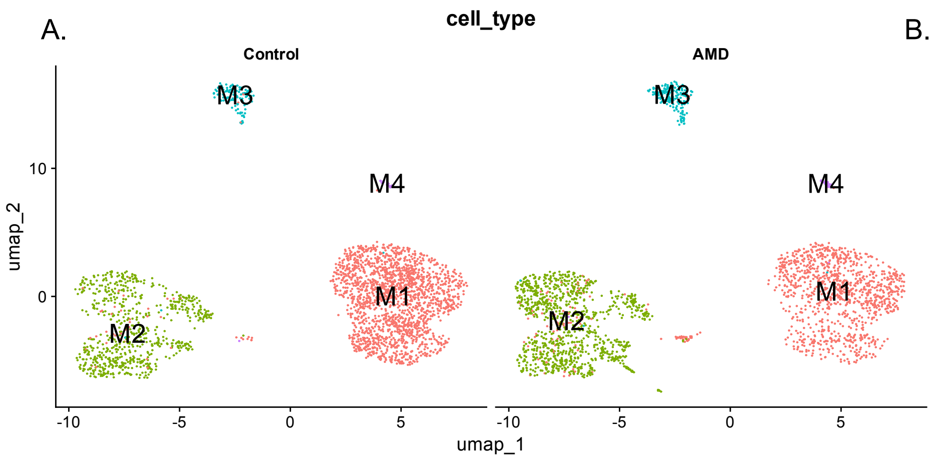

B.

| | Control | AMD |
|---|---|---|
| M1 | 1877 | 1170 |
| M2 | 699 | 1026 |
| M3 | 111 | 174 |
| M4 | 20 | 26 |

**Supplementary Figure 18**

(A) UMAP showing the projected cell clusters when subdividing Microglia cell cluster into smaller groups between the Control (13) and AMD (17) samples.
(B) Table showing the number of cells in each cluster in controls and AMD.

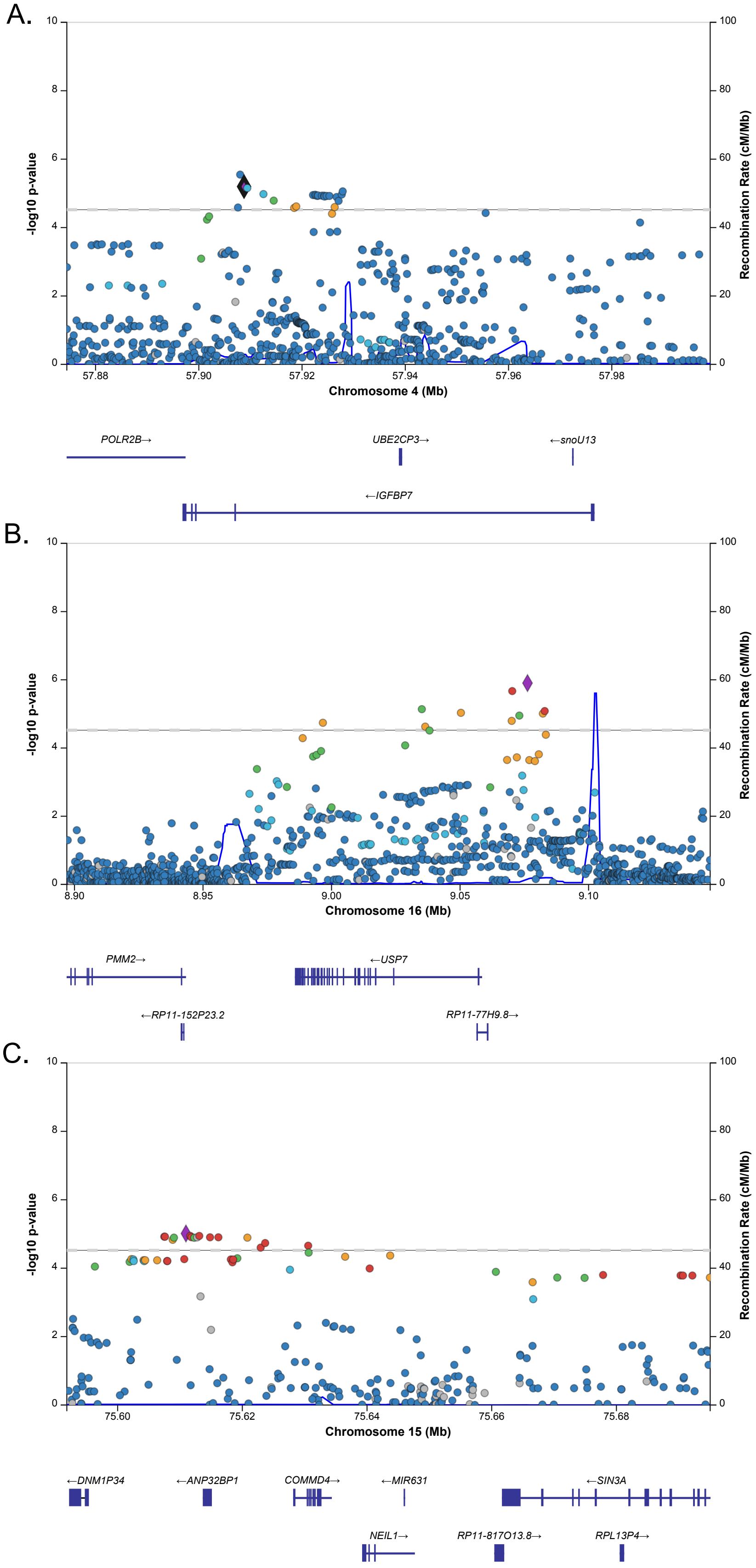

A.
-log10 p-value
Recombination Rate (cM/Mb)
Chromosome 4 (Mb)
57.88
57.90
57.92
57.94
57.96
57.98
POLR2B→
UBE2CP3→
←snoU13
←IGFBP7
B.
-log10 p-value
Recombination Rate (cM/Mb)
Chromosome 16 (Mb)
8.90
8.95
9.00
9.05
9.10
PMM2→
←USP7
←RP11-152P23.2
RP11-77H9.8→
C.
-log10 p-value
Recombination Rate (cM/Mb)
Chromosome 15 (Mb)
75.60
75.62
75.64
75.66
75.68
←DNM1P34
←ANP32BP1
COMMD4→
←MIR631
←SIN3A
NEIL1→
RP11-817O13.8→
RPL13P4→

**Supplementary Figure 19**

(A) Locus Zoom plot showing suggestive association (p-value < 5 x$10^{-5}$) for variants around IGFBP7 with AMD.
(B) Locus Zoom plot showing suggestive association (p-value < 5 x$10^{-5}$) for variants around USP7 with AMD.
(C) Locus Zoom plot showing suggestive association (p-value < 5 x$10^{-5}$) for variants around NEIL1 with AMD.

| | Cell Type | Cell Count | Percentage |
|---|---|---|---|
| UK (3 donors) | Amacrine | 24 | 0.22% |
| | Astrocyte | 27 | 0.24% |
| | Cones | 577 | 5.22% |
| | Horizontal | 46 | 0.42% |
| | Microglia | 107 | 0.97% |
| | Muller Glia | 452 | 4.09% |
| | Off-Cone Bipolar | 460 | 4.16% |
| | On-Cone Bipolar | 319 | 2.88% |
| | RGC | 221 | 2.00% |
| | Rod Bipolar | 945 | 8.54% |
| | Rods | 7883 | 71.27% |
| | | 11061 | |
| | Cell Type | Cell Count | Percentage |
| Mullins 1 sample | Amacrine | 22 | 1.07% |
| | Astrocyte | 27 | 1.31% |
| | Cones | 0 | 0.00% |
| | Horizontal | 15 | 0.73% |
| | Microglia | 12 | 0.58% |
| | Muller Glia | 326 | 15.80% |
| | Off-Cone Bipolar | 188 | 9.11% |
| | On-Cone Bipolar | 47 | 2.28% |
| | RGC | 0 | 0.00% |
| | Rod Bipolar | 231 | 11.20% |
| | Rods | 1195 | 57.93% |
| | | 2063 | |
| | Cell Type | Cell Count | Percentage |
| Scheetz 2 samples | Amacrine | 0 | 0.00% |
| | Astrocyte | 18 | 0.43% |
| | Cones | 0 | 0.00% |
| | Horizontal | 0 | 0.00% |
| | Microglia | 97 | 2.32% |
| | Muller Glia | 1925 | 46.13% |
| | Off-Cone Bipolar | 276 | 6.61% |
| | On-Cone Bipolar | 98 | 2.35% |
| | RGC | 0 | 0.00% |
| | Rod Bipolar | 630 | 15.10% |
| | Rods | 1129 | 27.05% |
| | | 4173 | |

**Supplementary Table 1**

Table showing the number of cells in each Cell Type after annotation, divided by datasets.

### AAK 81 Genes

| gene | avg_FC | p_val_adj | cluster |
|---|---|---|---|
| C3 | 2.65 | 5.16E-09 | Microglia |
| HLA-DPA1 | 2.25 | 1.38E-07 | Microglia |
| RNASET2 | 3.03 | 5.65E-07 | Microglia |
| HLA-DRA | 2.13 | 5.01E-05 | Microglia |
| SLC9A9 | 0.78 | 5.73E-05 | Microglia |
| CYTH4 | 0.67 | 0.0013697 | Microglia |
| TRIM22 | 1.58 | 0.00263638 | Microglia |
| HLA-B | 1.72 | 0.00756009 | Microglia |
| PLCG2 | 2.02 | 2.47E-16 | Astrocyte |
| SERPINA5 | 2.64 | 5.04E-12 | Astrocyte |
| LINC01411 | 2.10 | 0.0005 | Astrocyte |
| F5 | 1.53 | 0.0216 | Astrocyte |
| PLCG2 | 2.01 | 3.92E-60 | Müller Glia |
| MOXD1 | 1.46 | 1.89E-28 | Müller Glia |
| HLA-B | 1.46 | 5.60E-20 | Müller Glia |
| B2M | 1.36 | 3.86E-09 | Müller Glia |
| PLCG2 | 1.33 | 2.51E-50 | RGC |
| B2M | 1.31 | 1.13E-11 | RGC |
| HLA-B | 1.31 | 2.52E-09 | RGC |

### High Confidence Genes

| gene | avg_FC | p_val_adj | cell type |
|---|---|---|---|
| APOE | 4.969302 | 9.00E-22 | microglia |
| PARP12 | 1.45821731 | 0.01078179 | microglia |
| VEGFA | 1.70775175 | 0.04219704 | microglia |
| APOE | 3.23072995 | 3.58E-20 | astrocyte |
| RLBP1 | 1.32582703 | 4.70E-05 | astrocyte |
| TRPM1 | 1.89937481 | 0.00930651 | astrocyte |
| APOE | 2.94736455 | 3.10E-217 | mullerglia |
| PILRB | 2.16412782 | 1.59E-80 | mullerglia |
| VEGFA | 1.5157062 | 5.13E-35 | mullerglia |
| HTRA1 | 1.2928636 | 3.30E-13 | mullerglia |
| TRPM1 | 1.58460941 | 2.01E-10 | mullerglia |
| PILRB | 2.26332398 | 4.48E-100 | RGC |
| BLOC1S1 | 2.05938625 | 6.35E-76 | RGC |
| APOE | 2.32537345 | 2.15E-60 | RGC |
| VEGFA | 1.36300085 | 8.74E-33 | RGC |

**Supplementary Table 2**

Tables showing the differential expression result of the 81 ML-genes and known AMD genes between Control and AMD samples in each cell type.

| Chr | Pos | SNP id | Gene | AMD-GWAS p-value |
|---|---|---|---|---|
| chr16 | 81885924 | rs4133124 | PLCG2 | 2.6E-06 |
| chr16 | 81885272 | rs12921746 | PLCG2 | 3.2E-05 |
| chr16 | 81890366 | rs4889424 | PLCG2 | 7.8E-05 |
| chr16 | 81890788 | rs7202169 | PLCG2 | 8.5E-05 |
| chr16 | 81874766 | rs7200783 | PLCG2 | 9.1E-05 |
| chr16 | 81890509 | rs4889426 | PLCG2 | 9.4E-05 |
| chr4 | 57041844 | rs1718877 | IGFBP7 | 2.8E-06 |
| chr4 | 57042596 | rs34857305 | IGFBP7 | 6.3E-06 |
| chr4 | 57043222 | rs1277272 | IGFBP7 | 7E-06 |
| chr4 | 57056375 | rs10031585 | IGFBP7 | 1.1E-05 |
| chr4 | 57055954 | rs4561981 | IGFBP7 | 1.1E-05 |
| chr4 | 57056533 | rs1713960 | IGFBP7 | 1.2E-05 |
| chr4 | 57056936 | rs1713958 | IGFBP7 | 1.2E-05 |

**Supplementary Table 3**
Variants around *PLCG2* and *IGFBP7* that were tested for function in luciferase activity.
